# Mapping domains of early-life determinants of future multimorbidity across three UK longitudinal cohort studies

**DOI:** 10.1101/2024.02.01.24301771

**Authors:** S Stannard, A Berrington, SDS Fraser, S Paranjothy, RB Hoyle, RK Owen, A Akbari, M Shiranirad, R Chiovoloni, NA Alwan

**Affiliations:** School of Primary Care, Population Sciences and Medical Education, Faculty of Medicine, University of Southampton, Southampton, United Kingdom; School of Economic, Social and Political Sciences, University of Southampton, Southampton, United Kingdom; School of Medicine, Medical Sciences and Nutrition, University of Aberdeen□; School of Mathematical Sciences, University of Southampton, Southampton, United Kingdom; Population Data Science, Swansea University Medical School, Faculty of Medicine, Health & Life Science, Swansea University, Swansea, United Kingdom; University Hospital Southampton NHS Foundation Trust, Southampton, United Kingdom

## Abstract

Many studies use a reductionist approach to isolate the influence of one factor in childhood on multimorbidity rather than consider the combined effect of wider determinants. We explored how potential multiple early-life determinants of multimorbidity can be characterised across three UK cohort studies.

We used the National Child Development Study (NCDS), the 1970 British Cohort Study (BCS70), and the Aberdeen Children of the 1950s Study (ACONF) to identified early-life variables that fit into 12 domains of early-life determinants of multimorbidity. Variables were assigned into 12 domains; principal component analysis reduced the dimensionality of the data and structured variables into subgroups.

The data audit identified 7 domains in ACONF, 10 domains in NCDS and 12 domains in BCS70. Components included maternal fertility histories within the prenatal, antenatal and birth domain, long-term illnesses within the child health domain, educational ability within the child education and health literacy domain, ethnicity within the demography domain, parental health behaviours within the transgenerational domain, housing within the socioeconomic domain and parental-child interactions within the parental-family domain.

Conceptualising the risk of future multimorbidity as lifecourse domains composed of multiple factors can help challenge the existing understanding of disease aetiology and develop new ideas for prevention of multimorbidity.

## Introduction

A substantial body of evidence points to the very early part of life being crucial in determining health in childhood and in later years. Exposures in childhood, such as parental social class, family income, parental employment, poor health, and adverse childhood experiences are associated with the risk of multimorbidity in adulthood, defined as the co-existence of two or more chronic conditions [1-5]. However, it is increasingly recognised that health is multidetermined and its risk factors are complex. Yet, many studies use a reductionist approach to isolate the influence of one specific factor on health outcomes rather than consider the combined effect of wider determinants from across the lifecourse [6]. This is in part because data limitations often preclude the analysis of multiple variables simultaneously, and because this multidetermined nature of health is challenging to characterise, map and explore. Additionally, authors sometimes explicitly reduce the number of variables chosen for analysis to reduce statistical complexity and computation that can result from highly correlated variables. Despite this, we know that risk factors for disease are interlinked and often cluster together [7], so separating them into independent domains is challenging but should be considered. Therefore, researchers should examine clusters of risk, not just individual risk factors, which is the more traditional epidemiological approach.

Developing methods for capturing, mapping, and exploring how multimorbidity risk factors cluster in domains, as opposed to analysing individual exposures, can help develop more effective public health solutions. Identifying populations at higher risk and understanding how to address the clustering of vulnerabilities to future ill health requires approaches to data analyses that take account of the complexities of how risk is shaped and amplified across the lifecourse. In earlier work [8], we conceptually identified 12 domains of early life risk factors as being important for multimorbidity risk. These domains were developed from a review of existing research evidence and policy, and co-produced with public involvement. This conceptualisation was built on the concept of lifecourse epidemiology, defined by Kuh and Ben-Shlomo [9] as ‘the study of long-term effects on later health or disease risk of physical or social exposures during gestation, childhood, adolescence, young adulthood and later adult life’.

The 12 domains outlined in Figure 1 included: *Domain 1: Prenatal, antenatal, neonatal and birth (from conception to the first month of life)* that focused on the period from preconception to the onset of labour, the circumstances and outcomes surrounding a birth, and the period immediately following birth. *Domain 2: Adverse childhood experiences (ACEs)* described negative experiences or events such as abuse, neglect, domestic violence, parental substance abuse, parental death, parental separation, and parental incarceration. *Domain 3: Child health* considered the health of a child from birth to age 18. *Domain 4: Developmental attributes and behaviour* (under the age of 18) focused on the developmental markers of children relating to cognition, coordination, personality types and behavioural traits, and included diagnosed neurodevelopmental conditions. *Domain 5: Child education* related to the process of learning and educational achievement, especially in educational settings, and the knowledge an individual gains from these educational institutions. *Domain 6: Demographics* referred to factors that described the size, structure, and distribution of populations. *Domain 7: Transgenerational impact of parental health, behaviours and education* referred to factors that can be transmitted across generations. *Domain 8: Socioeconomic factors* included factors concerned with the interaction of social and economic issues. *Domain 9:* The *parental and family environment* incorporated parental-child interactions and the interaction between children and the primary care giver, parenting styles, parental beliefs, attitudes and discipline, and wider family factors such as kin networks. *Domain 10: Neighbourhood, the physical environmental and health care systems* incorporated external factors relating to neighbourhoods and the physical environments. *Domain 11: Health behaviours and diet* incorporated common health behaviours and diet. *Domain 12: Religion, spirituality and wider culture* combined the role of religion, spirituality and wider cultural norms and attitudes on influencing health, health literacy, health behaviours and health care decisions. We conceptually identified these 12 domains of future multimorbidity risk a priori of how they are represented within data. Therefore, our aim in this paper is to explore how these 12 childhood domains of future multimorbidity risk are characterised and represented within available data in the UK.

**Figure 1:**
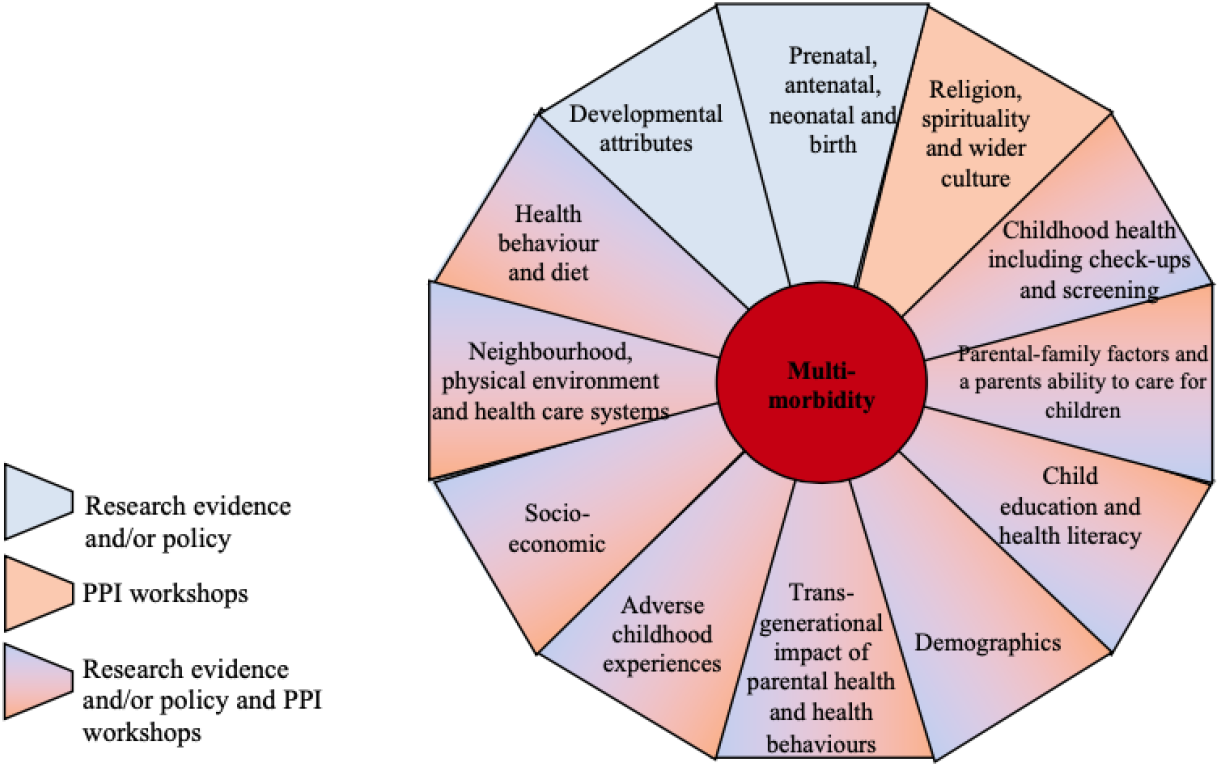
The 12 domains of early-life risk factors identified from the literature, policy and PPI contributions [8].

This work is conducted as part of the Multidisciplinary Ecosystem to study Lifecourse Determinants and Prevention of Early-onset Burdensome Multimorbidity (MELD-B) project [10], which aims to use an epidemiological and an artificial intelligence enhanced analysis of birth cohort and Electronic Healthcare data sources to identify lifecourse time periods and targets for the prevention of early-onset, burdensome multiple long-term conditions.

## Methods

### Data sources

To map the conceptualised domains to the available and relevant variables, it was important we consider data sources that captured a wide array of biological, social, environmental, and family variables from across childhood, and that follow up respondents across adulthood. Therefore, we focused on three UK longitudinal cohort studies. The *Aberdeen Children of the 1950s (ACONF)* includes children born in Aberdeen, Scotland, between 1950 and 1956; in total there are 12,150 cohort members, and participants were traced in their forties (2002) and linked to hospital and mental health admissions, maternity records, cancer registers, and death records [11]. The main ‘reading survey’ comprised of reading and maths test taken at school, and cohort members were linked to other school and birth records. A ‘family survey’ was administered randomly to 1 in 5 mothers of the cohort members regarding a range of topics, including the child’s medical history, mother’s attitude towards their child’s education and their aspirations for them, leisure activities of the child and parent, and housing conditions and social background of the parents [11]. The *National Child Development Study (NCDS)* [12] follows all children born in England, Scotland and Wales in one week in 1958, and includes 17,415 cohort members. To date, there have been 11 sweeps of data collection – 4 in childhood and 6 in adulthood, and a biomedical sweep of data collected was conducted via a healthcare visitor at age 42. The *1970 British Cohort Study (BCS70)* [13] follows 17,096 cohort members born in England, Scotland, Wales, and Northern Ireland in one week in 1970. To date, there have been 10 sweeps of data collection – 4 in childhood and 6 in adulthood, and a biomedical sweep of data collected was conducted via a healthcare visitor at age 46. All three data sources have collected information on social, economic, biological, and environmental factors at various time points in childhood.

### Analyses

The first stage involved a manual data audit that mapped the 12 conceptualised domains to the available and relevant variables in childhood across the three data sources. Initially, we reviewed all variables recorded in the BCS70 and NCDS at ages 10 and 11, and ACONF at ages 7-12. Each of the variables were grouped into the domain they best represented. We acknowledge overlaps between domains, and that some variables could have been recorded across multiple domains. However, we chose not to duplicate variables across domains to prevent multiple counting, instead we included a variable in the domain the research team mutually felt it best represented.

Variables were excluded from the data audit if they did not represent any of the 12 domains i.e., they were not relevant to any of the 12 domains. Given that the BCS70 and NCDS collected additional data at birth, age 5 and 16 (BCS70) and age 7 and 16 (NCDS), we expanded the data landscape audit to include variables reported in these additional sweeps to supplement domains that were either poorly represented or were of poor data quality such as high levels of missing, at age 10 (BCS70) and 11 (NCDS). A list of all the variables identified from the data landscape audit are included in Supplementary Table 1-3.

After allocating all the variables identified from the data audit to a specific domain, we conducted an exploratory analysis to understand the relationship between the variables within a domain. This exploratory analysis included producing Pearson correlation coefficients and Principal Component Analysis (PCA).

Firstly, Pearson correlation coefficients were used to identify any highly correlated variables within a domain; this was an important step for reducing multicollinearity prior to any future regression modelling. We defined highly correlated variables as those with a correlation coefficient greater than 0.7 [14]. Any correlated variables were initially flagged prior to the PCA analysis. A strength of the PCA analysis is that it identified highly correlated variables and transformed these variables into a smaller set of variables, called principal components. However, if highly correlated variables identified from the Pearson correlation coefficients remained after the PCA analysis, we retained the flag in the data audit. This flag was retained to allow researcher to make their own decision on which variables to retain prior to any further modelling; we suggest that decisions on which variables to retain should be based on data quality (i.e., level of missing), the strength of the theoretical and/or conceptual reasoning for retaining a variable within a domain, and the specific research questions being addressed.

Secondly, PCA analysis was performed to help reduce the dimensionality of the data by categorising each of the 12 domains into mutually exclusive PCA components based on similar characteristics [15]. Individual PCA components were scaled to have mean zero, the data was automatically standardised to have unit variance, and PCA components within each domain were identified if they had an eigenvalue of above one [16,17]. If domains had multiple PCA components with an eigenvalue above one, we selected the top four components. Finally, we identified individual variables important for a PCA component if they had a component score above 0.3 [18,19]. We defined important PCA components as the component that contributed the greatest proportion of the overall variance for each domain and a single data source. Therefore, importance indicates a combined variable (PCA component) that provides the most variation within a single data source for a single domain. The PCA components within each domain were given a descriptive name chosen by the research team that summarised all the variables within a component with a component score above 0.3. Given PCA analysis does not perform well on categorical data, where appropriate, we used multiple factor analysis as an extension of PCA analysis to deal with categorical variables [20]. Analysis was conducted using STATA 17.0, and figures were created using online diagram software ‘drawio’ [21].

## Results

### Data landscape audit and domain data mapping

#### ACONF

After conducting the data landscape audit in the ACONF dataset, 74 variables were identified that represented 7 of the conceptualised domains (i.e., demographic, socioeconomic, developmental attributes, childhood health, education and health literacy, antenatal, neonatal and birth and neighbourhood domains) (Figure 2). Several domains were represented by variables recorded in the family survey - a subset of the full ACONF sample (2,208/12,150 cohort members). Therefore, a decision was made for those domains where data were present in two surveys (i.e., the demographic, socioeconomic, and developmental attributes), to only include data from the full ACONF sample. The only exception was the childhood health domain, which could only be captured in the family survey. The data audit did not find enough data to sufficiently represent the ACE domain; religion, spirituality, wider culture domain; parental-family factors domain; transgenerational domain and health behaviours and diet domain.

**Figure 2.**
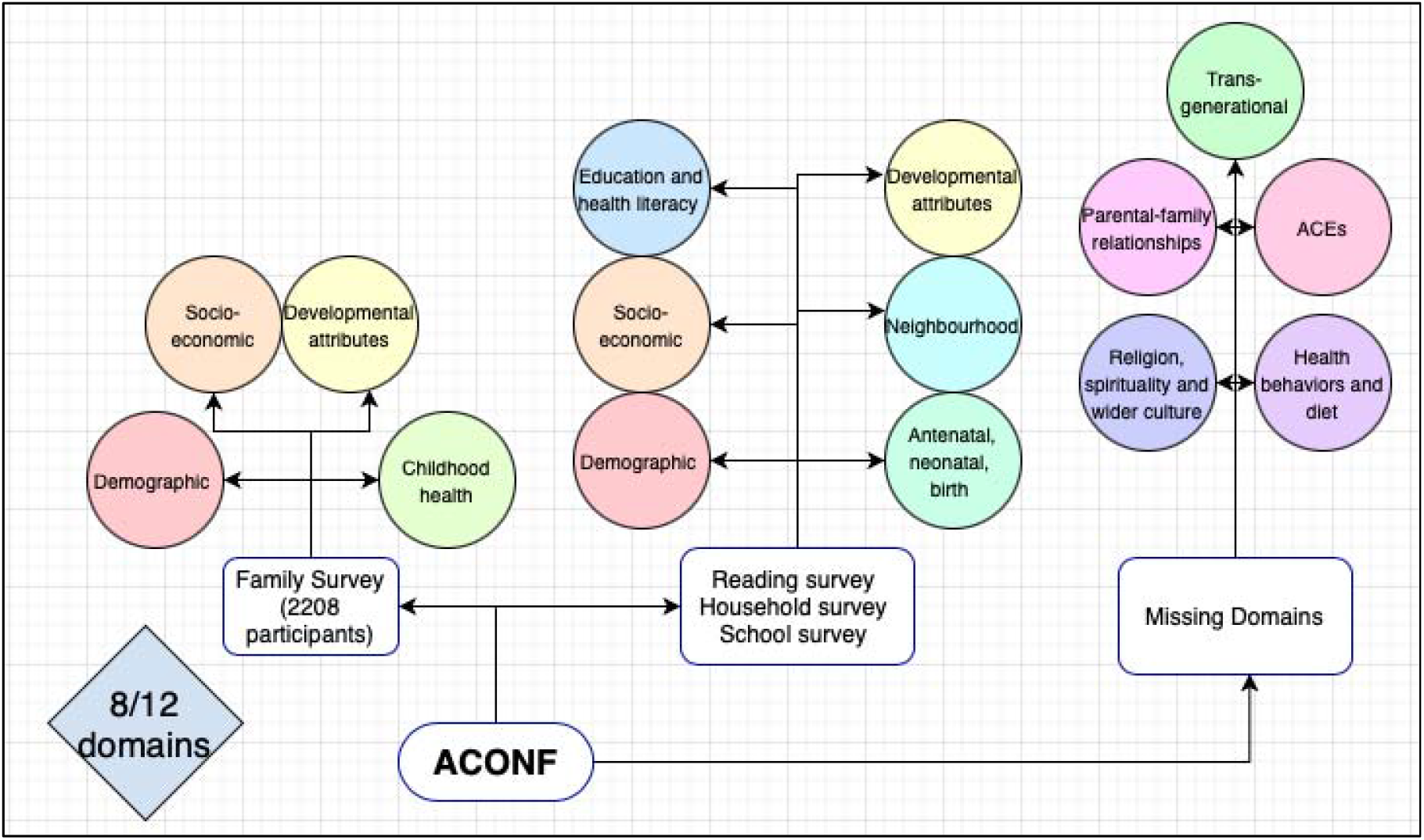
Data landscape audit mapping the available and relevant variables in ACONF to the conceptualised domain.

#### NCDS

As shown in Figure 3, after conducting the data landscape audit in the NCDS dataset, 143 variables were identified that represented 10 of the conceptualised domains (i.e., demographic, socioeconomic, developmental attributes, childhood health, transgenerational, antenatal, neonatal and birth, neighbourhood, health behaviours and diet, parental-family relations and education and health literacy domains) (Figure 3). As demonstrated, supplementary sweeps of data collection at birth, age 7 and age 16 were utilised to supplement the antenatal, neonatal and birth domain (birth sweep) the health behaviour and diet domain, and the transgenerational and neighbourhood domains (ages 7 and 16 sweeps), as these domains were not represented at age 11. The data audit in the NCDS dataset did not discover enough data to accurately represent the ACE domain or religion, spirituality, or wider culture domain.

**Figure 3.**
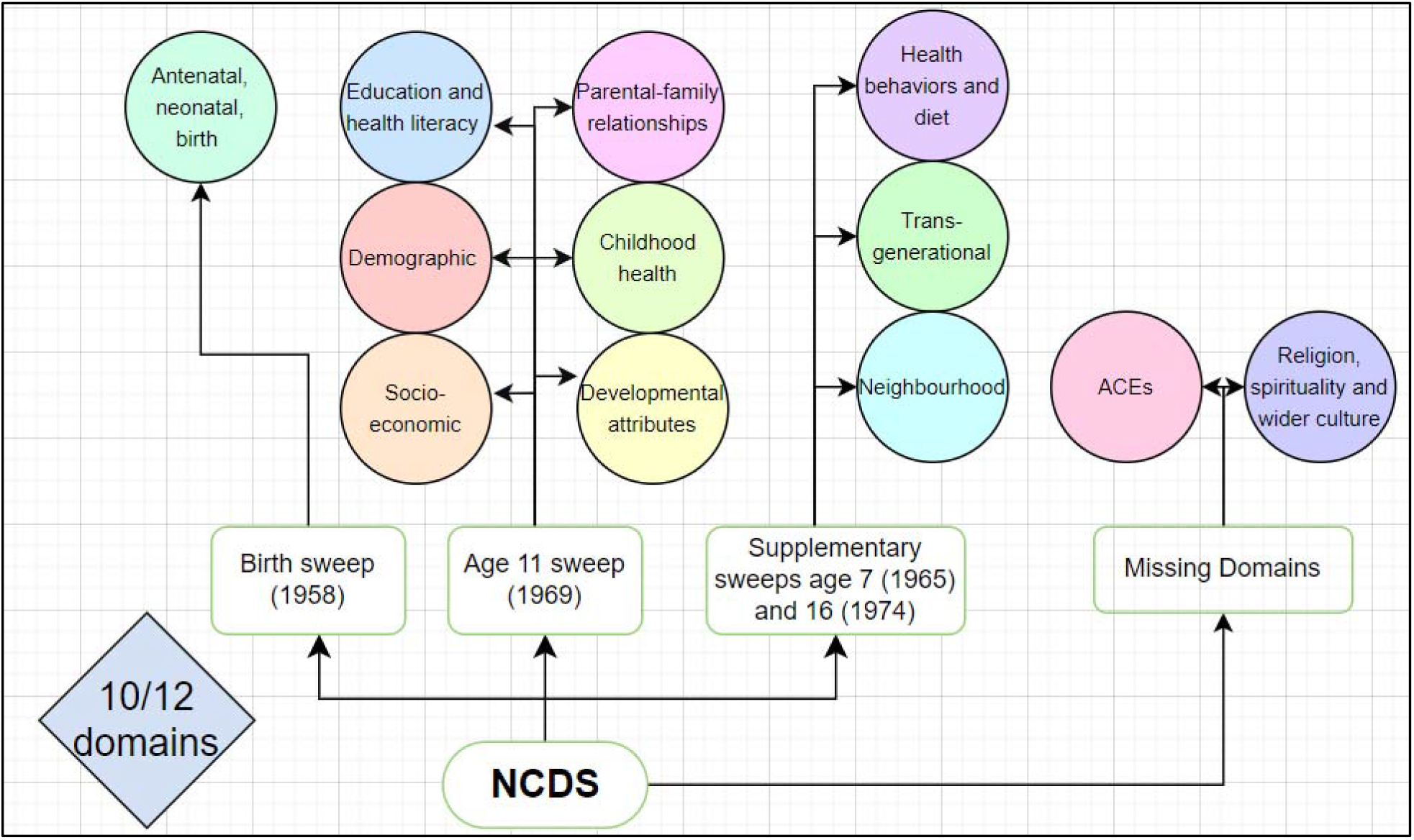
Data landscape audit mapping the available and relevant variables in NCDS to the conceptualised domains.

#### BCS70

As shown in Figure 4 after conducting the data landscape audit in the BCS70 dataset, 289 variables were identified that represented all 12 of the conceptualised domains. As demonstrated, supplementary sweeps of data collection at birth, age 5, and age 16 were utilised to supplement the antenatal, neonatal and birth domain (birth sweep), health behaviour and diet domain, and ACE domain (ages 5 and 16 sweep), as these domains were not well represented at age 10 in the BCS70. We also supplemented the age 10 transgenerational domain and neighbourhood domain with data recorded at ages 5 and 16, given these domains had high levels of missing at age 10.

**Figure 4.**
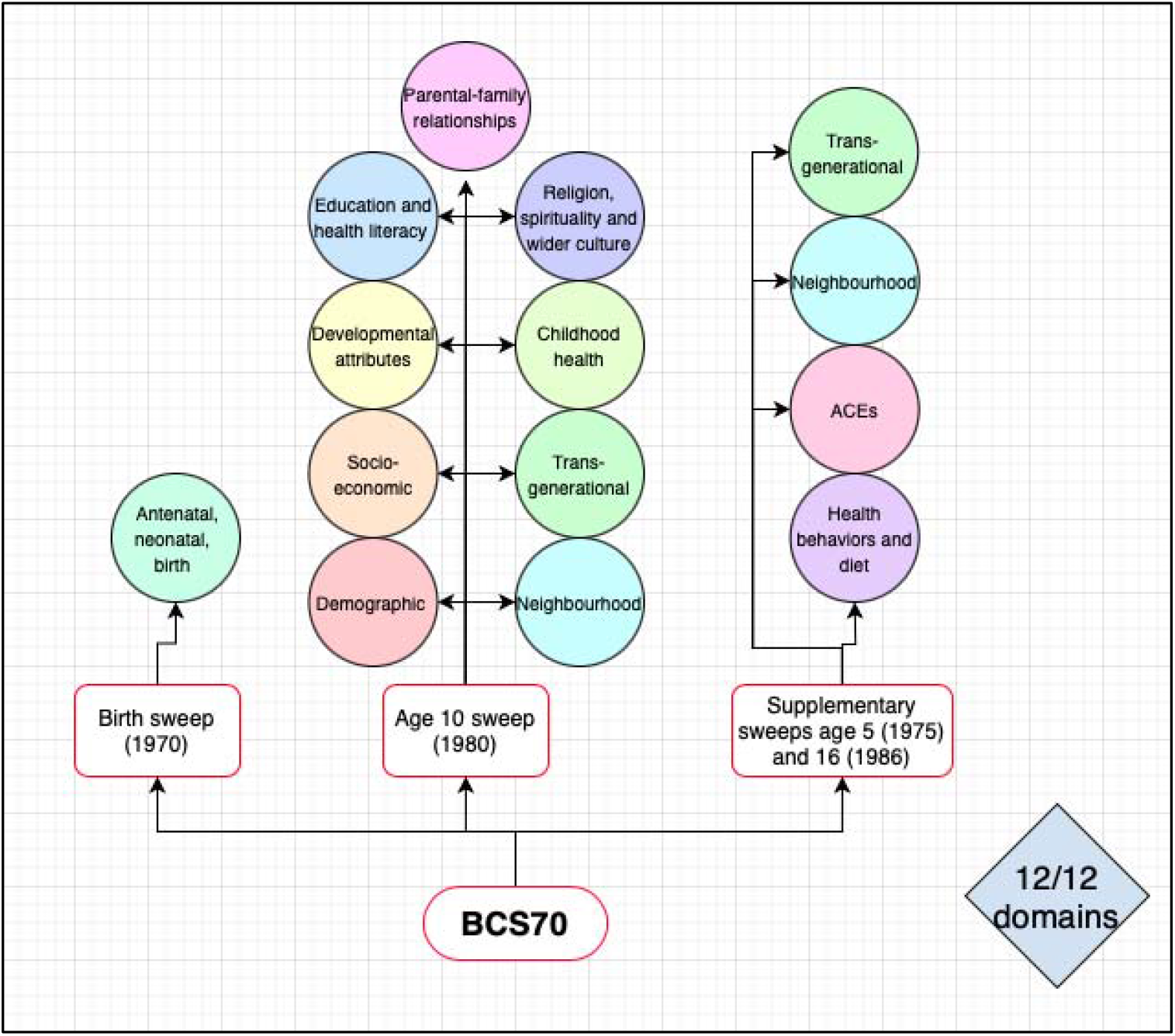
Data landscape audit mapping the available and relevant variables in BCS70 to the conceptualised domains.

### Correlation coefficients and PCA analysis

The full results from the PCA analysis are included in Supplementary Figure 1-3. The component that contributed the greatest proportion of the overall variance for each domain (i.e., component 1) are highlighted in Table 1. Utilising PCA analysis to identify mutually exclusive groups reduced the dimensionality of the ACONF variables from 74 to 41. Important within domain components included a physical grade component that contributed 15% of the variance to domain 1 (prenatal, antenatal, neonatal and birth). This component included variables relating to maternal age, ‘mother physical grade’ (condition of mother at birth) and ‘child physical grade’ (condition of baby at birth). A behavioural component that contributed 56% of the variance to domain 4 (developmental attributes) and included variables on ‘neurotic/anti-social rating’ and ‘total score scale b’ (a measure of child behaviour). An IQ and parental education component that contributed 30% of the variance to domain 5 (education) that incorporated variables focusing on school mean IQ and mother’s further education and father’s further education. A family size component that contributed 24% of the variance to domain 6 (demographic) included variables relating to position of index child and family size. A housing component that contributed 35% of the variance to domain 8 (socioeconomic) incorporated variables relating to person per room, housing tenure and housing area. An amenity in area component contributed 35% of the variance to domain 10 (neighbourhood, environment, and health care systems) and included variables relating to access to cold water, a bath and a shared WC, and whether a house is being rented from the council.

**Table 1.**
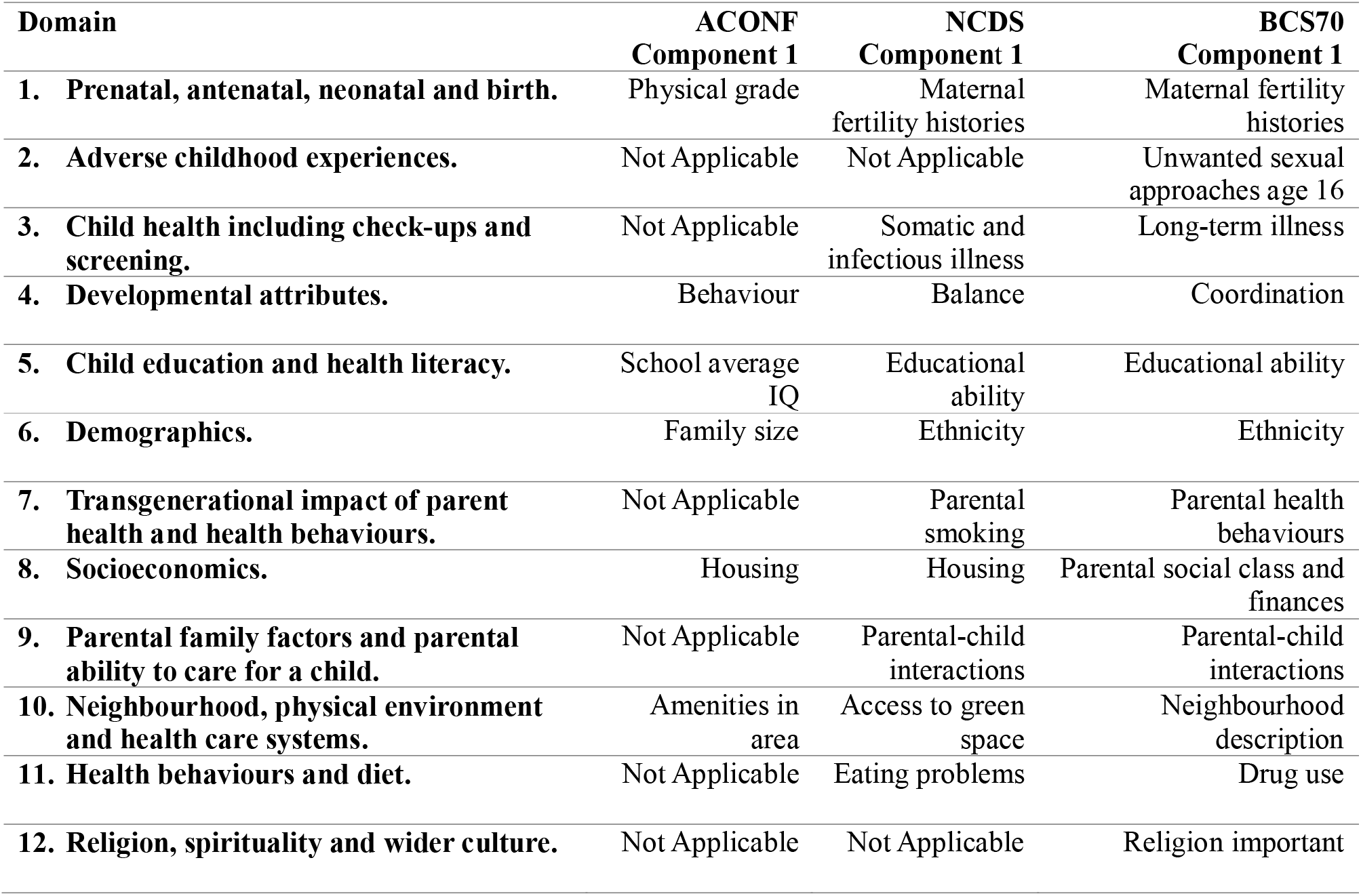
The component for each domain that contributed the greatest proportion of the overall variance (i.e., Component 1), for all three data sources.

Utilising PCA reduced the dimensionality of the NCDS data recorded at birth and ages 7, 11 and 16 from 142 variables to 73 variables. Important within domain components included maternal fertility histories that contributed 14% of the variance to domain 1 (prenatal, antenatal, neonatal and birth), and variables within this component included maternal age, parity, and birth spacing. A ‘somatic’ and ‘infectious’ illness component that contributed 7% of the variance to domain 3 (child health), that incorporated variables relating to ‘somatic symptoms’ and ‘infectious illnesses’ reported in childhood. A balance component that contributed 25% of the variance to domain 4 (developmental attributes), and included variables assessing walking in a straight line, standing on left and right leg, and balancing heel to toe. An educational ability component contributed 59% of the variance to domain 5 (education), and incorporated variables focusing on general knowledge, number, book, oral, math and general ability tests, and reading comprehension. An ethnicity component contributed 19% of the variance to domain 6 (demographic), and included variables relating to language spoken in home, mother’s ethnicity, and father’s ethnicity.

Other important components in the NCDS included parental smoking that contributed 25% of the variance to domain 7 (transgenerational), and included variables related to parental smoking. A housing component contributed 20% of the variance to domain 8 (socioeconomic), and incorporated variables relating to household number, sharing a bedroom and number of persons per room. A parental-child interactions component contributed 18% of the variance to domain 9 (parental-family factors) and included variables relating to the mother and father going on walks with the child and father helping with managing a child. An access to green space component contributed 26% of the variance to domain 10 (neighbourhood, environment, and health care systems), and included variables relating to access to play areas, access to public parks and access to recreational grounds. Finally, an eating problems component contributed 24% of the variance to domain 11 (health behaviours) and included variables relating to any eating disorders and the type of eating disorders.

Utilising PCA analysis in the BCS70 and on data recorded at birth and ages 5, 10 and 16, reduced the dimensionality of the dataset from 289 variables to 149 variables. Important within domain components included a maternal fertility histories component that contributed 16% of the variance to domain 1 (prenatal, antenatal, neonatal and birth), and variables within this component included maternal age, parity, and number of previous pregnancies. An unwanted sexual approaches (age 16) component contributed 11% of the variance to domain 2 (adverse childhood experiences) and included variables relating to the number of ‘unwanted sexual approaches in the last year’. A long-term illness component contributed 11% of the variance to domain 3 (child health), and included abnormal gastrointestinal finding, abnormal neurological findings, abnormal endocrine findings, and abnormal mental handicap findings. A coordination component contributed 18% of the variance to domain 4 (developmental attributes), and included variables focussing on hand coordination, being clumsy at games, difficulty picking up a objects and difficulty kicking a ball. An educational ability component contributed 23% of the variance to domain 5 (education) and incorporated variables focusing on reading and math tests, estimated reading ages, difficulty reading and writing, and reading ability. An ethnicity component contributed 35% of the variance to domain 6 (demographic) and included variables concerning child’s ethnicity, mother’s ethnicity and father’s ethnicity.

Other important components in the BCS70 included a parental health behaviours component that contributed 8% of the variance to domain 7 (transgenerational), and included variables related to maternal smoking, father’s smoking, mother’s healthy lifestyle and father’s healthy lifestyle. A parental social class and finance component contributed 18% of the variance to domain 8 (socioeconomic), and incorporated variables relating to parental social class, van/car ownership, family income and living on a council estate. A parental-child interactions component contributed 10% of the variance to domain 9 (parental-family factors) and encompassed variables relating to the family doing activities together (walks/outings/meals/holiday/shopping/restaurants) and families chatting for at least 5 minutes per day. A neighbourhood description component contributed 21% of the variance to domain 10 (neighbourhood, environment, and health care systems), and included variables concerning noisy neighbourhood, teenagers on streets, ‘drunks’ on the street and rubbish on the street. A drug use component contributed 10% of the variance to domain 11 (health behaviours), and combined variables relating to drug use (heroin/semeron/cocaine/downers/uppers). Finally, a religion important component contributed 22% of the variance to domain 12 (religion, spirituality, and culture), and incorporated variables in relation to the religion a person was born into, time spent on religion, if religion was important and if religious views are misguided.

## Discussion

In this paper we conducted a data landscape audit across three UK longitudinal cohort studies to map early life variables against a conceptual framework of lifecourse determinants of multimorbidity. We categorised three cohort datasets into mutually exclusive components based on similar within domain characteristics. Eight domains were characterised by 74 variables in ACONF recorded when participants were aged 7-12, ten domains were characterised by 143 variables in the NCDS recorded at the birth of the participant or at ages 7, 11 and 16, and twelve domains were characterised by 289 variables in the BCS70 recorded at the birth of the participant or at ages 5, 10 and 16. PCA analysis reduced the dimensionality of ACONF variables from 74 to 41, from 143 to 73 in the NCDS, and from 289 to 149 in the BCS70.

The data audit successfully mapped all 12 conceptualised domains [8] to the available and relevant variables across the three datasets. For some domains we have partial coverage across the data sources - religion, spirituality, and wider culture domain (BCS70), adverse childhood experience domain (BCS70), child health including check-ups and screening domain (NCDS and BCS70) and the parental family factors and parental ability to care for a child domain (NCDS and BCS70). The remaining domains are represented across all ages in childhood within all three data sources.

Important components based on similar within domain characteristics across the three datasets included maternal fertility histories within the prenatal, antenatal and birth domain (NCDS and BCS70), long-term or ‘somatic’ symptoms within the child health, including check-up and screening domain (NCDS and BCS70), IQ or educational ability within the child education and health literacy domain (ACONF, NCDS and BCS70). Other important components included ethnicity within the demography domain (NCDS and BCS70), parental health behaviours within the transgenerational impact of parent health and health behaviours domain (NCDS and BCS70), housing within the socioeconomic domain (ACONF and NCDS) and parental-child interaction within the parental family factors and parental ability to care for a child domain (NCDS and BCS70).

We know that single exposures in childhood are associated with the risk of multimorbidity in adulthood [1-5]. However, it is complex and difficult to explore the combined action of health determinants across childhood both because data limitations often preclude the analysis of multiple determinants, and because such combinations are difficult to characterise, map and explore in terms of their components and timing. As a result, most studies have used a reductionist approach to isolate the influence of a specific factor on health outcomes rather than consider the multidetermined nature of combined factors [6]. We have demonstrated that the auditing, characterisation and mapping of potential early life determinants of future health outcomes can be achieved if multiple large scale longitudinal studies are used. We have demonstrated the strength in using multiple prospective studies to consider the potential long-term effects of combined domains of risk relating to social, economic, and environmental exposures during early life. Although we note that this study was lacking consistency regarding the uniformity of variables across data sources.

Acting on these wider combinations of determinants from across childhood will not just improve health outcomes but have further intermediate benefits, such as narrowing social, environmental and health inequalities in childhood that may be on the pathway to health outcomes in adulthood. Conceptually multiple combined risk factors might lead to greater risk of single disease outcomes such as cardiovascular disease [22,23], however the concept of developing methods for capturing the multidimensional nature of risk of multiple disease outcomes such as multimorbidity is a field of growing interest and research [24,25]. This mapping and characterisation of variables that relate to early life risk domains of multimorbidity can provide a step towards understanding and promoting population-level action to prevent or delay the burden of multimorbidity. This work also supports the drive within the UK healthcare system about shifting towards a more preventive model of health. The Department of Health and Social Care [26] policy paper on transforming the public health system highlighted the need to focus on prevention and the wider determinants of health, and the 2018 paper on the Public Health Priorities in Scotland [27] included the need to invest early in young people’s future as the best form of prevention. We therefore see our research as an extension to these discussions and a contribution to this emerging field, as we have demonstrated that through the effective utilisation and evaluation of birth cohort datasets, we were able to successfully audit and map how domains for prevention of multiple long-term conditions can be represented within data.

## Future research

Given we have conceptually [8] and now methodologically captured potential domains of early life risk factors of future multimorbidity risk, our next will look at quantifying the association between these domains and/or potential interaction between domains and their impact on risk of developing multimorbidity within the MELD-B project [10]. Additionally, we may look to explore the relationship across domains paying particular attention to how domains cluster together. The data audit is also useful for other researchers who wish to use cohort data. We have demonstrated and documented the array of important early childhood variables within a domain, and these could be used to address a range of research questions and topics, not just related to health.

## Strengths and limitations

The three longitudinal cohort studies provide some of the richest and most in-depth data in the UK and allowed us to capture a wide array of biological, social, environmental, and family variables from across the whole of childhood. The same level of information would not have been available from electronic health care records in either primary or secondary care. Despite this, there were conceptualised domains of multimorbidity risk outlined in our previous work [8] such as religion, spirituality and wider culture domain (NCDS & ACONF) the ACE domain (NCDS & ACONF), child health including check-ups and screening domain (ACONF), parental family factors and parental ability to care for a child domain (ACONF) and health behaviour and diet domain (ACONF) that we were unable to analyse across all three datasets. This was either because they were of poor data quality or only reported in one of the cohorts. It is also important to note that variables were not identical across data sources, meaning direct comparisons should be interpreted with caution. The three cohort studies are also representative of a cohort of children born in 1950-1956 (ACONF), 1958 (NCDS) and 1970 (BCS70), and as such these data sources largely lack ethnic diversity and do not reflect the population of the UK today. A further limitation of using historical data sources was that some of the variables should be interpreted in the historical context in which they were recorded, newer surveys such as the Millennium Cohort Study may have improved variables that better represent some of the domains.

We acknowledge that PCA analysis represents a simple and straightforward method to reduce the dimensionality of these data. More sophisticated methods could be used such as latent class analysis or clustering methods, some of which will be explored in future work as parts of the MELD-B project [10]. It is important to note that we define important combined variables (PCA components) as those that provide the most variation within a single data source for a single domain. It was beyond the scope of this paper to explore a) whether the variable, component or domain we have identified are important predictors of future long-term conditions or b) whether the component identified provides the most variation within a domain across the population as a whole / multiple datasets, because this depends very much on which variables were measured and available to compare.

## Conclusion

We have demonstrated that if multiple large scale longitudinal studies are used, it is possible to move beyond a reductionist approach that isolates the influence of a specific factor on health to consider combined risk factors. Developing such approaches for capturing, mapping, and exploring the combined effects of early life risk factors for health as opposed to individual exposures can help to challenge traditional epidemiological approach to the aetiology of disease, and develop new ideas and solutions for the prevention of ill health. Further research should build on the data audit to explore the relationship between the domains we have identified and risk of developing multimorbidity.

## Conflicts of Interest statement

RKO is a member of the National Institute for Health and Care Excellence (NICE) Technology Appraisal Committee, member of the NICE Decision Support Unit (DSU), and associate member of the NICE Technical Support Unit (TSU). She has served as a paid consultant to the pharmaceutical industry and international reimbursement agencies, providing unrelated methodological advice. She reports teaching fees from the Association of British Pharmaceutical Industry (ABPI). RBH is a member of the Scientific Board of the Smith Institute for Industrial Mathematics and System Engineering.

## Funding statement

This work is part of the <u>multidisciplinary ecosystem to study lifecourse determinants and prevention of early-onset burdensome multimorbidity (MELD-B)</u> project which is supported by the National Institute for Health Research (NIHR203988). The views expressed are those of the authors and not necessarily those of the NIHR or the Department of Health and Social Care.

## Authors Contributions

S.F., N.A., R.H., S.P., R.O., A.A., S.S. and A.B. contributed to the conceptualisation of the MELD-B project. S.S., N.A. and S.F. obtained the datasets. S.S. and N.A. led the design and planning of the analyses. All authors contributed to the conceptualisation of the analyses. S.S. performed the statistical analysis and prepared all figures and graphs. S.S., N.A. and A.B. produced the initial draft of the manuscript. All authors were involved in editing and reviewing the manuscript, and approved the final manuscript. S.S., N.A. and S.F. take responsibility for the data and research governance.

## Acknowledgements

We would like to acknowledge all other members of the MELD-B Consortium: Michael Boniface, Emilia Holland, Ruben Sanchez-Garcia, Zlatko Zlatev, Jessica Enright, Martin Gulliford, Alex Dregan, Paul Smart, Nic Fair, Gareth Giles, Aiysha Qureshi, Chandni Jacob, Jim McMahon, Rosie Martin, Jack Welch, Rebecca Longley, Peter Williams, Sara Macdonald, Mark Ashworth, Kelly Cheung, Nick Francis, Frances Mair, Rita Rajababoo, Saroj Parekh, Lynn Laidlaw, Samina Begum Sally Dace, Christine Kemp-Philp, Heather Parsons, Becky Wilkinson, Louise Coutts.

We acknowledge the support of the DaSH who provided secure data storage and management. We thank the participants of the NCDS, BCS70 and ACONF cohort studies.

## Data Availability Statement

The BCS70 and NCDS datasets generated and analysed in the current study are available from the UK Data Archive repository (available here: http://www.cls.ioe.ac.uk/page.aspx?&sitesectionid=795). ACONF data can be accessed via the Grampian Data Safe Haven by accredited researchers with appropriate research governance approvals (available here: https://www.abdn.ac.uk/iahs/facilities/grampian-data-safe-haven.phpen <u>(abdn.ac.uk)</u>.

## Notes

### Funding Statement

This work stems from reflections within the multidisciplinary ecosystem to study lifecourse determinants and prevention of early-onset burdensome multimorbidity (MELD-B) project which is supported by the National Institute for Health Research (NIHR203988). The views expressed are those of the authors and not necessarily those of the NIHR or the Department of Health and Social Care.

### Author Declarations

The study is conducted in accordance with the UK Policy Framework for Health and Social Care Research. Ethics approval has been obtained from the University of Southampton Faculty of Medicine Ethics committee (ERGO II Reference 66810).

## References

1. Harper, S., Lynch, J., & Smith, G. D. Social determinants and the decline of cardiovascular diseases: understanding the links. Annu Rev Public Health. 32:39–69 (2011).

2. Humphreys, J., Jameson, K., Cooper, C., & Dennison, E. Early-life predictors of future multimorbidity: results from the Hertfordshire Cohort. Age Ageing. 47(3):474–478 (2018).

3. Johnston, M. C., Black, C., Mercer, S. W., Prescott, G. J., & Crilly, M. A. Impact of educational attainment on the association between social class at birth and multimorbidity in middle age in the Aberdeen Children of the 1950s cohort study. BMJ Open.;9(1):e024048 (2019).

4. Gondek, D., Bann, D., Brown, M., Hamer, M., Sullivan, A., & Ploubidis, G. B. Prevalence and early-life determinants of mid-life multimorbidity: evidence from the 1970 British birth cohort. BMC Public Health. 21(1):1319 (2021).

5. Zhao, M., He, X., Li, T., Shao, H., Huo, Q., & Li, Y. Early-Life Factors and Multimorbidity Risk Later in Older Age: Evidence Based on CHARLS. Gerontology. 69 (11): 1347–1357 (2023).

6. Alvidrez, J., Greenwood, G. L., Johnson, T. L., & Parker, K. L. Intersectionality in Public Health Research: A View From the National Institutes of Health. Am J public health. 111(1), 95–97 (2021).

7. Birch, J., Petty, R., Hooper, L., Bauld, L., Rosenberg, G., & Vohra, J. Clustering of behavioural risk factors for health in UK adults in 2016: a cross-sectional survey. J Public Health. 41(3), e226–e236 (2019).

8. Stannard, S., et al., A conceptual framework for characterising lifecourse determinants of multiple long-term condition multimorbidity. J Multimorb Comorb. (2023).

9. Kuh, D., & Ben-Shlomo, Y., eds. A life course approach to chronic disease epidemiology; tracing the origins of ill-health from early to adult life. 2nd edn. (Oxford: Oxford University Press 1997).

10. Fraser S.D., et al., Multidisciplinary ecosystem to study lifecourse determinants and prevention of early-onset burdensome multimorbidity (MELD-B) – protocol for a research collaboration. J Multimorb Comorb. (2023).

11. Batty, G. D., et al., The Aberdeen Children of the 1950s cohort study: background, methods, and follow-up information on a new resource for the study of life-course and intergenerational effects on health. Paediatr Perinat Epidemiol. 18: 221–239 (2004).

12. Power, C., & Elliott, J. Cohort profile: 1958 British birth cohort (National Child Development Study). International J Epidemiol. 35(1), 34–41 (2009).

13. Sullivan, A., Brown, M., Hamer, M., & Ploubidis., G. Cohort Profile Update: The 1970 British Cohort Study (BCS70). International J Epidemiol. dyac148 (2022).

14. Boslaugh, S., & Watters, P. A. Inferential Statistics in Statistics in a Nutshell: A Desktop Quick Reference. 125–151. (Sebastopol, CA: O’Reilly Media, 2008).

15. Zota, A. R., & VanNoy, B. N. Integrating Intersectionality Into the Exposome Paradigm: A Novel Approach to Racial Inequities in Uterine Fibroids. Am J Public Health. 111(1), 104–109 (2021).

16. Boehmke, B., & Greenwell, B. M. Principal Components Analysis in Hands-On Machine Learning with R (1st ed.). (Chapman and Hall/CRC, New York, 2019).

17. Stata Manuals. Pca – Principal component analysis. https://www.stata.com/manuals/mvpca.pdf (2023).

18. Grimm, L. G., & Yarnold, P. R. Reading and understanding multivariate statistics. (Washington, DC: American Psychological Association, 1995).

19. Hair, J.F., Tatham, R.L., Anderson, R.E. & Black, W. Multivariate data analysis. (Fifth Ed.) (Prentice-Hall:London, 1998).

20. Abdi, H., Williams, L. J., & Valentin, D. Multiple factor analysis: principal component analysis for multitable and multiblock data sets. WIREs Comp Stat. 5 149–179, 2013.

21. Drawio https://www.drawio.com/ (2023)

22. Framingham. Hard Coronary Heart Disease (10-year risk). Hard Coronary Heart Disease (10-year risk) | Framingham Heart Study (2001).

23. QRISK®3 calculator. ClinRisk. https://www.qrisk.org (2023).

24. Wister, A., et al,. Development and validation of a multi-domain multimorbidity resilience index for an older population: Results from the baseline Canadian Longitudinal Study on Aging. BMC Geriatrics. 18(1):1–13, (2018).

25. Wister, A., Klasa, K., & Linkov, L. A Unified Model of Resilience and Aging: Applications to COVID-19. Public Health Front. 886: 1–14, (2022).

26. Department of Health and Social Care. Transforming the public health system: reforming the public health system for the challenged of our time. Transforming the public health system: reforming the public health system for the challenges of our times - GOV.UK (http://www.gov.uk), (2021).

27. Public Health Scotland. Scotland’s public health priority. Public Health Priorities for Scotland (http://www.gov.scot), (2018).

