## Supplementary Tables 1-3 for "Mapping domains of early-life determinants of future multimorbidity across three UK longitudinal cohort studies"

**Supplementary Table 1. Variables identified from the data audit in BCS70**

| Variable | Description | Sweep of data collection | Supplementary variable | Domain |
|---|---|---|---|---|
| TeenMum | EVER A TEENAGE MOTHER (BD1AGEFB GROUPED) | Birth | No | Prenatal, antenatal, neonatal and birth |
| MatAge | AGE OF MOTHER AT CM'S BIRTH (FROM S1 VAR A0005A/S2 VAR E008) | Birth | No | Prenatal, antenatal, neonatal and birth |
| MatSmoke | SMOKING DURING PREGNANCY | Birth | No | Prenatal, antenatal, neonatal and birth |
| NoPrevPreg | NO OF PREVIOUS PREGNANCIES (ENTRIES ON FILE) | Birth | No | Prenatal, antenatal, neonatal and birth |
| Parity | PARITY | Birth | No | Prenatal, antenatal, neonatal and birth |
| NoAnteVisits | NUMBER OF ANTENATAL VISITS | Birth | No | Prenatal, antenatal, neonatal and birth |
| AnteInpVisit | OCCURRENCE OF ANTENATAL INPATIENT CARE | Birth | No | Prenatal, antenatal, neonatal and birth |
| PlaceDel | PLACE OF DELIVERY | Birth | No | Prenatal, antenatal, neonatal and birth |
| LabDur | DURATION 1ST STAGE OF LABOUR (HOURS | Birth | No | Prenatal, antenatal, neonatal and birth |
| MethodDel | METHOD OF DELIVERY | Birth | No | Prenatal, antenatal, neonatal and birth |
| Birthweight | BIRTHWEIGHT OF BABY IN GRAMS | Birth | No | Prenatal, antenatal, neonatal and birth |
| AnyOp | WERE ANY OPERATIONS PERFORMED | Birth | No | Prenatal, antenatal, neonatal and birth |
| Resus | BABY RESUSCITATION | Birth | No | Prenatal, antenatal, neonatal and birth |
| CongAbno | BORN WITH CONGENITAL ABNORMALITY | Birth | No | Prenatal, antenatal, neonatal and birth |
| Illness | BABY ILL AT BIRTH | Birth | No | Prenatal, antenatal, neonatal and birth |
| EverCongAbnor | CHILD EVER HAD ANY ABNORMALITY | 10 | No | Child health including check-ups and screening |
| EverBronchitis | CHILD EVER HAD BRONCHITIS | 10 | No | Child health including check-ups and screening |
| EverMeasles | EVER HAD MEASLES | 10 | No | Child health including check-ups and screening |
| EverMumps | EVER HAD MUMPS | 10 | No | Child health including check-ups and screening |
| EverWhooping | EVER HAD WHOOPING COUGH | 10 | No | Child health including check-ups and screening |

| | | | | |
|---|---|---|---|---|
| EverChickenPox | EVER HAD CHICKEN POX | 10 | No | Child health including check-ups and screening |
| EverMeningitis | EVER HAD MENINGITIS | 10 | No | Child health including check-ups and screening |
| EverAcuteFever | EVER HAD OTHER ACUTE FEVER | 10 | No | Child health including check-ups and screening |
| EverTonsillectomy | EVER HAD TONSILLECTOMY | 10 | No | Child health including check-ups and screening |
| EverHerniaOp | EVER HAD HERNIA OPERATION | 10 | No | Child health including check-ups and screening |
| EverAppendicetomy | EVER HAD APPENDICECTOMY | 10 | No | Child health including check-ups and screening |
| EverOppSquint | EVER HAD OPERATION FOR SQUINT | 10 | No | Child health including check-ups and screening |
| EverVaccine | CHILD EVER HAD IMMUNISATION, VACCINATION | 10 | No | Child health including check-ups and screening |
| VacDiphtheria | VACCINATED AGAINST DIPHTHERIA | 10 | No | Child health including check-ups and screening |
| VacWhoopingCough | VACCINATED AGAINST WHOOPING COUGH | 10 | No | Child health including check-ups and screening |
| VacTetanus | VACCINATED AGAINST TETANUS | 10 | No | Child health including check-ups and screening |
| VacSmallpox | VACCINATED AGAINST SMALLPOX | 10 | No | Child health including check-ups and screening |
| VacPolio | VACCINATED AGAINST POLIOMYELITIS | 10 | No | Child health including check-ups and screening |
| VacMeasles | VACCINATED AGAINST MEASLES | 10 | No | Child health including check-ups and screening |
| VacTB | VACCINATED AGAINST TB (BCG) | 10 | No | Child health including check-ups and screening |
| VacOther | VACCINATED AGAINST OTHER | 10 | No | Child health including check-ups and screening |
| EverAdmHosp | HAS CHILD EVER BEEN ADMITTED TO HOSPITAL | 10 | No | Child health including check-ups and screening |
| TotalHospAdm | TOTAL NUMBER OF HOSPITAL ADMISSIONS | 10 | No | Child health including check-ups and screening |
| Outpatients | HAS CHILD EVER ATTENDED OUTPATIENTS? | 10 | No | Child health including check-ups and screening |

| | | | | |
|---|---|---|---|---|
| TotalAccidents | TOTAL NUMBER OF ACCIDENTS | 10 | No | Child health including check-ups and screening |
| ChildSeenPsychiatrist | CHILD EVER BEEN TO CHILD PSYCHIATRIST | 10 | No | Child health including check-ups and screening |
| EmotionalBehaProb | HAS CHILD EVER HAD EMOTION,BEHAV. PROBS | 10 | No | Child health including check-ups and screening |
| IllnessHandicap | ANY EVIDENCE OF ANY ILLNESS/HANDICAP ETC | 10 | No | Child health including check-ups and screening |
| SoreThroat12m | RECURRENT SORE THROATS, PAST 12 MONTHS | 10 | No | Child health including check-ups and screening |
| MiddleEar12m | MIDDLE EAR INFECTION, PAST 12 MONTHS | 10 | No | Child health including check-ups and screening |
| HearingLoss12m | HEARING LOSS, PAST 12 MONTHS | 10 | No | Child health including check-ups and screening |
| Eczema12m | ECZEMA, PAST 12 MONTHS | 10 | No | Child health including check-ups and screening |
| HayFever12m | HAY FEVER, PAST 12 MONTHS | 10 | No | Child health including check-ups and screening |
| Asthma12m | ASTHMA, PAST 12 MONTHS | 10 | No | Child health including check-ups and screening |
| Wheezy12m | WHEEZY BRONCHITIS, PAST 12 MONTHS | 10 | No | Child health including check-ups and screening |
| Bronchitis12m | BRONCHITIS, PAST 12 MONTHS | 10 | No | Child health including check-ups and screening |
| Pneumonia12m | PNEUMONIA, PAST 12 MONTHS | 10 | No | Child health including check-ups and screening |
| HeartCond12m | PATHOLOGICAL HEART COND, PAST 12 MONTHS | 10 | No | Child health including check-ups and screening |
| AbdomPain12m | RECURRENT ABDOM PAIN, PAST 12 MONTHS | 10 | No | Child health including check-ups and screening |
| Hernia12m | INGUINAL HERNIA, PAST 12 MONTHS | 10 | No | Child health including check-ups and screening |
| UrinaryInf12m | URINARY INFECTION, PAST 12 MONTHS | 10 | No | Child health including check-ups and screening |
| OtherIllness12m | OTHER ILLNESSES, PAST 12 MONTHS | 10 | No | Child health including check-ups and screening |
| SystolicBP | BLOOD PRESSURE SYSTOLIC | 10 | No | Child health including check-ups and screening |

| | | | | |
|---|---|---|---|---|
| DiastolicBP | BLOOD PRESSURE DIASTOLIC | 10 | No | Child health including check-ups and screening |
| AbnFancialGeneral | ABNORMAL FINDINGS A) FACIAL AND GENERAL | 10 | No | Child health including check-ups and screening |
| AbnSkin | ABNORMAL FINDINGS B) SKIN CONDITION | 10 | No | Child health including check-ups and screening |
| AbnENT | ABNORMAL FINDINGS C) ENT CONDITION | 10 | No | Child health including check-ups and screening |
| AbnUpperResp | ABNORMAL FINDINGS D) UPPER RESPIRATORY | 10 | No | Child health including check-ups and screening |
| AbnLowerResp | ABNORMAL FINDINGS E) LOWER RESPIRATORY | 10 | No | Child health including check-ups and screening |
| AbnCardiovasc | ABNORMAL FINDINGS F) CARDIOVASCULAR | 10 | No | Child health including check-ups and screening |
| AbnGastro | ABNORMAL FINDINGS G) GASTROINTESTINAL | 10 | No | Child health including check-ups and screening |
| AbnAbdoOther | ABNORMAL FINDINGS H) OTHER ABDOMINAL | 10 | No | Child health including check-ups and screening |
| AbnUrogenital | ABNORMAL FINDINGS I) UROGENITAL | 10 | No | Child health including check-ups and screening |
| AbnNeuro | ABNORMAL FINDINGS J) NEUROLOGICAL | 10 | No | Child health including check-ups and screening |
| AbnMusculo | ABNORMAL FINDINGS K) MUSCULO SKELETAL | 10 | No | Child health including check-ups and screening |
| AbnEndocrine | ABNORMAL FINDINGS L) ENDOCRINE | 10 | No | Child health including check-ups and screening |
| AbnBloodLymp | ABNORMAL FINDINGS M) BLOOD, LYMPHATIC | 10 | No | Child health including check-ups and screening |
| AbnMentalHandicap | ABNORMAL FINDINGS N) MENTAL HANDICAP | 10 | No | Child health including check-ups and screening |
| AbnBehEmot | ABNORMAL FINDINGS O) BEHAVIOUR/EMOTIONAL | 10 | No | Child health including check-ups and screening |
| CongAbnor | ANY EVIDENCE OF CONGENITAL ABNORMALITY | 10 | No | Child health including check-ups and screening |
| ChildBuild | DESCRIPTION OF CHILD'S BUILD | 10 | No | Child health including check-ups and screening |
| ConvReason | CONVULSION REASON | 10 | No | Child health including check-ups and screening |

| | | | | |
|---|---|---|---|---|
| BMI | BMI | 10 | No | Child health including check-ups and screening |
| RutterBeh | TOTAL RUTTER BEHAVIOUR SCORE - GROUPED | 10 | No | Developmental attributes |
| EmotionProb | HAS CHILD EVER HAD EMOTION,BEHAV. PROBS | 10 | No | Developmental attributes |
| IllHandicap | ANY EVIDENCE OF ANY ILLNESS/HANDICAP ETC | 10 | No | Developmental attributes |
| ThrowCatchBall | THROWING BALL AND CATCHING | 10 | No | Developmental attributes |
| ThrowCatchClap | THROW BALL, CATCH BOTH HANDS, NO. CLAPS | 10 | No | Developmental attributes |
| StandRightLeg | STANDING ON RIGHT LEG, DID FOOT MOVE | 10 | No | Developmental attributes |
| StandLeftLeg | STANDING ON LEFT LEG, DID FOOT MOVE | 10 | No | Developmental attributes |
| WalkBackwards | WALKING BACKWARDS, NO. STEPS | 10 | No | Developmental attributes |
| Clumsy | DESCRIPTION OF CHILD'S COORDINATION | 10 | No | Developmental attributes |
| HandCoord | SCALE WORKS DEFTLY WITH HANDS | 10 | No | Developmental attributes |
| Temper | SCALE DISPLAYS OUTBURSTS OF TEMPER | 10 | No | Developmental attributes |
| ClumsyGame | SCALE CLUMSY AT GAMES | 10 | No | Developmental attributes |
| DiffKickBall | SCALE DIFFICULTY KICKING BALL | 10 | No | Developmental attributes |
| DiffPickingUp | SCALE DIFFICULTY PICKING UP SMALL OBJECTS | 10 | No | Developmental attributes |
| EdinbRead | EDINBURGH READING TEST SCORE (I3003 TO I3069) | 10 | No | Education and health literacy |
| EstReadAge | ESTIMATED READING AGE AT AGE 10 (DERIVED FROM ZB10READ AND AVERAGE AGE) | 10 | No | Education and health literacy |
| FriendlyMath | FRIENDLY MATHS TEST SCORE (I2504 TO I2575) | 10 | No | Education and health literacy |
| DiffMaths | DIFFICULTY WITH SCHOOL SUBS MATHS | 10 | No | Education and health literacy |
| DiffRead | READING | 10 | No | Education and health literacy |
| DiffWrite | WRITING | 10 | No | Education and health literacy |
| GenKnowledge | CHILD'S GENERAL KNOWLEDGE | 10 | No | Education and health literacy |
| IndepWork | DOES CHILD WORK INDEPENDENTLY | 10 | No | Education and health literacy |
| AbilityMaths | ABILITY IN MATHS | 10 | No | Education and health literacy |
| AbilityRead | ABILITY IN READING | 10 | No | Education and health literacy |
| AbilitySpell | ABILITY IN SPELLING | 10 | No | Education and health literacy |

| | | | | |
|---|---|---|---|---|
| AbilityWrite | ABILITY IN CREATIVE WRITING | 10 | No | Education and health literacy |
| GoodSpell | GOOD AT SPELLING | 10 | No | Education and health literacy |
| TestGuess | TESTS ARE A LOT OF GUESS WORK | 10 | No | Education and health literacy |
| FatherQual | FATHERS ED QUALIFICATION | 10 | No | Education and health literacy |
| MotherQual | MOTHER ED QUALIFICATION | 10 | No | Education and health literacy |
| Region | STANDARD REGION OF RESIDENCE | 10 | No | Demographics |
| Sex | CHILD'S SEX | 10 | No | Demographics |
| HHNumber | NUMBER OF PERSONS IN HOUSEHOLD | 10 | No | Demographics |
| Parentalsep | CHILD LIVED WITH SAME PARENTS SINCE BORN | 10 | No | Demographics/ACE |
| ParentalDeath | REASON: DEATH OF A PARENT | 10 | No | Demographics/ACE |
| ChildEthnicity | ETHNIC GROUP STUDY CHILD 1 | 10 | No | Demographics |
| MotherEthnicity | ETHNIC GROUP MOTHER 1 | 10 | No | Demographics |
| FatherEthnicity | ETHNIC GROUP FATHER 1 | 10 | No | Demographics |
| IllMother | SINCE CHILD 5YR ILLNESS IN FAMILY MOTHER | 10 | No | Transgenerational impact of parent health and health behaviours |
| IllFather | SINCE CHILD 5YR ILLNESS IN FAMILY FATHER | 10 | No | Transgenerational impact of parent health and health behaviours |
| MotherDrinkEarly | DID MOTHER DRINK DURING PREGNANCY, EARLY | 10 | No | Transgenerational impact of parent health and health behaviours |
| MotherDrinkLate | DID MOTHER DRINK DURING PREGNANCY, LATER | 10 | No | Transgenerational impact of parent health and health behaviours |
| MotherCough | COUGH: MOTHER | 10 | No | Transgenerational impact of parent health and health behaviours |
| FatherCough | COUGH: FATHER | 10 | No | Transgenerational impact of parent health and health behaviours |
| FatherSmoke | MOTHER'S PRESENT SMOKING HABITS | 10 | No | Transgenerational impact of parent health and health behaviours |
| MotherSmoke | FATHER'S PRESENT SMOKING HABITS | 10 | No | Transgenerational impact of parent health and health behaviours |
| MotherBMI | MOTHER BMI | 10 | No | Transgenerational impact of parent health and health behaviours |
| FatherBMI | FATHER BMI | 10 | No | Transgenerational impact of parent health and health behaviours |
| SocialClass | SOCIAL CLASS FROM FATHERS OCCUP (OR MOTHERS IF MISSING) | 10 | No | Socioeconomics |

| | | | | |
|---|---|---|---|---|
| Benefits | RECEIVED STATE BENEFIT LAST 12 MONTHS? (C8.1 TO C8.11) | 10 | No | Socioeconomics |
| NumberHH | NUMBER OF PERSONS IN HOUSEHOLD | 10 | No | Socioeconomics |
| AccomType | WHAT ACCOMMODATION OCCUPIED BY FAMILY | 10 | No | Socioeconomics |
| HousingTenure | IS ACCOMMODATION OWNED OR RENTED? | 10 | No | Socioeconomics |
| UseBathroom | HAS FAMILY USE OF BATHROOM? | 10 | No | Socioeconomics |
| UseKitchen | HAS FAMILY USE OF KITCHEN? | 10 | No | Socioeconomics |
| Damp | IS ACCOMMODATION AFFECTED BY DAMP? | 10 | No | Socioeconomics |
| VanCar | DO YOU HAVE A CAR/VAN OF YOUR OWN | 10 | No | Socioeconomics |
| CouncilEstate | COUNCIL ESTATE | 10 | No | Socioeconomics |
| Income | AVERAGE INCOME | 10 | No | Socioeconomics |
| FatherEmploy | FATHER EMPLOYMENT STATUS | 10 | No | Socioeconomics |
| MotherEmploy | MOTHER EMPLOYMENT STATUS | 10 | No | Socioeconomics |
| MotherFig | RELATIONSHIP OF MOTHER FIGURE | 10 | No | Demographics/ACE |
| FatherFig | RELATIONSHIP OF FATHER FIGURE | 10 | No | Demographics/ACE |
| FatherManageChild | FATHER PLAYS ROLE IN MANAGING CHILD? | 10 | No | Parental family factors and parental ability to care for child |
| FamilyWalk | FAMILY ACTIVITIES A GO FOR WALKS | 10 | No | Parental family factors and parental ability to care for child |
| FamilyOutings | GO FOR OUTINGS TOGETHER | 10 | No | Parental family factors and parental ability to care for child |
| FamilyMeals | HAVE MEALS TOGETHER | 10 | No | Parental family factors and parental ability to care for child |
| FamilyHolidays | GO FOR HOLIDAYS TOGETHER | 10 | No | Parental family factors and parental ability to care for child |
| FamilyShopping | GO SHOPPING TOGETHER | 10 | No | Parental family factors and parental ability to care for child |
| FamilyChat | CHAT FOR AT LEAST 5 MINUTES | 10 | No | Parental family factors and parental ability to care for child |
| FamilyRestuarant | GO TO RESTAURANT TOGETHER | 10 | No | Parental family factors and parental ability to care for child |
| ParentMetTeach | PARENTS MET CHILD'S TEACHER | 10 | No | Parental family factors and parental ability to care for child |

| | | | | |
|---|---|---|---|---|
| ParentSchLeaveAge | AGE CHILD WILL LEAVE SCHOOL | 10 | No | Parental family factors and parental ability to care for child |
| ParentsDiscussTeach | PARENTS' DISCUSSIONS WITH TEACHER | 10 | No | Parental family factors and parental ability to care for child |
| MotherIntrEd | MOTHER'S INTEREST IN CHILD'S EDUCATION | 10 | No | Parental family factors and parental ability to care for child |
| FatherIntrEd | FATHER'S INTEREST IN CHILD'S EDUCATION | 10 | No | Parental family factors and parental ability to care for child |
| MotherHostile | -MOTHER'S ATTITUDE HOSTILE | 10 | No | Parental family factors and parental ability to care for child |
| MotherDismissive | MOTHER'S ATTITUDE DISMISSIVE | 10 | No | Parental family factors and parental ability to care for child |
| FatherHostile | FATHER'S ATTITUDE HOSTILE | 10 | No | Parental family factors and parental ability to care for child |
| FatherDismissive | FATHER'S ATTITUDE DISMISSIVE | 10 | No | Parental family factors and parental ability to care for child |
| ParentsHearIdeas | PARENTS LIKE TO HEAR ABOUT IDEAS | 10 | No | Parental family factors and parental ability to care for child |
| FoolishTalkParents | FOOLISH TALKING TO PARENTS | 10 | No | Parental family factors and parental ability to care for child |
| EverCare | EVER BEEN IN CARE | 10 | No | ACE |
| GoToPark | GOES TO PARK/PLAYGROUND | 10 | No | Neighbourhood, physical environments and health care systems |
| CloseTraffic | CLOSENESS OF TRAFFIC TO HOUSE | 10 | No | Neighbourhood, physical environments and health care systems |
| RuralArea | NEIGHBOURHOOD DESCRIPTION - RURAL | 10 | No | Neighbourhood, physical environments and health care systems |
| PercCatchClosPackedHouse | AREA: OF CLOSELY PACKED HOUSES | 10 | No | Neighbourhood, physical environments and health care systems |
| PercCatchCouncilEst | AREA: COUNCIL ESTATE OF HOUSES | 10 | No | Neighbourhood, physical environments and health care systems |
| PercCatchLessExpHouse | AREA: LESS EXPENSIVE PRIVATE | 10 | No | Neighbourhood, physical environments and health care systems |
| PercCatchWellSpacedHouse | AREA: WELL-SPACED HOUSES | 10 | No | Neighbourhood, physical environments |

| | | | | |
|---|---|---|---|---|
| | | | | and health care systems |
| PercCatchLargeHouse | AREA: LARGE HOUSES SET IN OWN GROUNDS | 10 | No | Neighbourhood, physical environments and health care systems |
| PercCatchRural | AREA: MAINLY RURAL | 10 | No | Neighbourhood, physical environments and health care systems |
| PercCatchOther | AREA: OTHER TYPE OF NEIGHBOURHOOD | 10 | No | Neighbourhood, physical environments and health care systems |
| DescribeTrafficSchCathment | DESCRIPTION OF TRAFFIC | 10 | No | Neighbourhood, physical environments and health care systems |
| Area | CATCHMENT AREA DESCRIPTION | 10 | No | Neighbourhood, physical environments and health care systems |
| SpareTimeSport | SPARE TIME ACTIVITIES: A SPORTS | 10 | No | Health behaviours and diet |
| SpareTimeWalks | GOES FOR WALKS | 10 | No | Health behaviours and diet |
| SpareTimeSwimming | GOES SWIMMING | 10 | No | Health behaviours and diet |
| FamilyWalks | FAMILY ACTIVITIES A GO FOR WALKS | 10 | No | Health behaviours and diet |
| SportOutsideClass | HOURS SPORT-OUTSIDE PERIODS | 10 | No | Health behaviours and diet |
| TriedCig | HAVE YOU EVER TRIED A CIGARETTE | 10 | No | Health behaviours and diet |
| ChocSweetsOften | HOW OFTEN DO YOU EAT CHOCOLATE/SWEETS? | 10 | No | Health behaviours and diet |
| VegRel | VEGETARIAN BECAUSE OF RELIGIOUS REASONS | 10 | No | Religion, spirtuality and wider culture |
| RelBornInto | WHAT RELIGION WERE YOU BORN INTO? | 10 | No | Religion, spirtuality and wider culture |
| RelImport | IS RELIGION IMPORTANT PART OF YOUR LIFE? | 10 | No | Religion, spirtuality and wider culture |
| RelLucky | RELIGIOUS PEOPLE LUCKY TO HAVE BELIEFS | 10 | No | Religion, spirtuality and wider culture |
| RelValuedSoc | RELIGIOUS PEOPLE VALUED MEMBERS OF SOC. | 10 | No | Religion, spirtuality and wider culture |
| RelMisguided | RELIGIOUS PEOPLE ARE MISGUIDED | 10 | No | Religion, spirtuality and wider culture |
| RelHelp | RELIGIOUS PEOPLE HELP YOU IF IN TROUBLE | 10 | No | Religion, spirtuality and wider culture |
| NoAlcohRel | DON'T DRINK ALCOHOL - RELIGION FORBIDS | 10 | No | Religion, spirtuality and wider culture |
| DietRel | IS SPECIAL DIET FOR RELIGION/CULTURE? | 10 | No | Religion, spirtuality and wider culture |
| TimeSpentRel | TOTAL TIME SPENT: RELIGIOUS ACTIVITY | 10 | No | Religion, spirtuality and wider culture |

| | | | | |
|---|---|---|---|---|
| Flag_Rel | FLAG FOR THOSE WHO REPORTED AT LEAST ONE QUESTION ON RELIGION | 10 | No | Religion, spirtuality and wider culture |
| MumAsthma5 | NAT. MUM - ASTHMA | 5 | Yes | Transgenerational impact of parent health and health behaviours |
| MumHayfever5 | NAT. MUM - HAYFEVER | 5 | Yes | Transgenerational impact of parent health and health behaviours |
| MumEczema5 | NAT. MUM - ECZEMA | 5 | Yes | Transgenerational impact of parent health and health behaviours |
| MumLateReader5 | NAT. MUM - LATE READER | 5 | Yes | Transgenerational impact of parent health and health behaviours |
| MumPoorReader5 | NAT. MUM - POOR READER | 5 | Yes | Transgenerational impact of parent health and health behaviours |
| MumConvulsions5 | NAT. MUM - CONVULSIONS | 5 | Yes | Transgenerational impact of parent health and health behaviours |
| MumLateSpeaker5 | NAT. MUM - LATE SPEAKER | 5 | Yes | Transgenerational impact of parent health and health behaviours |
| DadAsthma5 | NAT. DAD - ASTHMA | 5 | Yes | Transgenerational impact of parent health and health behaviours |
| DadHayfever5 | NAT. DAD - HAYFEVER | 5 | Yes | Transgenerational impact of parent health and health behaviours |
| DadEczema5 | NAT. DAD - ECZEMA | 5 | Yes | Transgenerational impact of parent health and health behaviours |
| DadLateReader5 | NAT. DAD - LATE READER | 5 | Yes | Transgenerational impact of parent health and health behaviours |
| DadPoorReader5 | NAT. DAD - POOR READER | 5 | Yes | Transgenerational impact of parent health and health behaviours |
| DadConvulsions5 | NAT. DAD - CONVULSIONS | 5 | Yes | Transgenerational impact of parent health and health behaviours |
| DadLateSpeaker5 | NAT. DAD - LATE SPEAKER | 5 | Yes | Transgenerational impact of parent health and health behaviours |
| MotherSmoke5 | SMOKING HABIT OF MOTHER | 5 | Yes | Transgenerational impact of parent health and health behaviours |
| FatherSmoke5 | SMOKING HABIT OF FATHER | 5 | Yes | Transgenerational impact of parent health and health behaviours |
| RelwithNeigh5 | RELATIONSHIP OF FAMILY WITH NEIGHBOURS | 5 | Yes | Neighbourhood, physical environments and health care systems |
| SocRatingNeigh5 | SOCIAL RATING OF NEIGHBOURHOOD | 5 | Yes | Neighbourhood, physical environments |

| | | | | |
|---|---|---|---|---|
| | | | | and health care systems |
| MaternalMalaise5 | TOTAL MALAISE SCORE - GROUPED IN STANDARD WAY | 5 | Yes | Transgenerational impact of parent health and health behaviours/ACE |
| NoHealthVisits5 | TOTAL HV VISITS SINCE BIRTH | 5 | Yes | Child health including check-ups and screening |
| NoCHCVisits5 | TOTAL CHC ATTENDANCES SINCE BIRTH | 5 | Yes | Child health including check-ups and screening |
| ScreeningPKU5 | ANY SCREENING FOR PKU LOGGED ON HV OR CHC RECORDS? | 5 | Yes | Child health including check-ups and screening |
| ScreeningCDH5 | ANY SCREENING FOR CDH (HIP) LOGGED ON HV OR CHC RECORDS? | 5 | Yes | Child health including check-ups and screening |
| ScreeningHearing5 | ANY SCREENING FOR HEARING LOGGED ON HV OR CHC RECORDS? | 5 | Yes | Child health including check-ups and screening |
| ScreeningSquint5 | ANY SCREENING FOR SQUINT LOGGED ON HV OR CHC RECORDS? | 5 | Yes | Child health including check-ups and screening |
| ScreeningVision5 | ANY SCREENING FOR VISION LOGGED ON HV OR CHC RECORDS? | 5 | Yes | Child health including check-ups and screening |
| TotalNoScreenings5 | TOTAL NUMBER OF SCREENINGS FOR GENERAL DEVELOPMENT, HEARING, VISION | 5 | Yes | Child health including check-ups and screening |
| AnyEatProb5 | EATING OR APPETITE PROBLEMS | 5 | Yes | Health behaviours and diet |
| EatProbUnderEat5 | NOT EATING ENOUGH | 5 | Yes | Health behaviours and diet |
| EatProbOvereating5 | OVEREATING | 5 | Yes | Health behaviours and diet |
| EatProbFaddiness5 | FADDINESS | 5 | Yes | Health behaviours and diet |
| OtherEatingProb5 | OTHER EATING PROBLEM | 5 | Yes | Health behaviours and diet |
| KeepSport16 | I AM KEEN ON SPORTS | 16 | Yes | Health behaviours and diet |
| PlaySportClub16 | LEISURE: PLAY SPORTS (AT CLUB/CENTRE ETC) | 16 | Yes | Health behaviours and diet |
| PlaySportStr~16 | LEISURE: PLAY SPORTS (STREET/PARK/ETC) | 16 | Yes | Health behaviours and diet |
| IntPeopleDif~16 | INTERESTED IN PEOPLE DIFF. RACE-RELIGION | 16 | Yes | Religion, spirtuality and wider culture |
| RelEdEssenti~16 | RELIGIOUS EDUC. IS ESSENTIAL IN SCHOOLS | 16 | Yes | Religion, spirtuality and wider culture |
| NoCigs16 | HOW MANY CIGARETTES DO YOU SMOKE A WEEK | 16 | Yes | Health behaviours and diet |
| SmokingHabits16 | DESCRIPTION SMOKING/NON SMOKING HABITS | 16 | Yes | Health behaviours and diet |

| | | | | |
|---|---|---|---|---|
| AlcoholPastW~16 | NUMBER OF DAYS HAD ALCOHOL PAST WEEK | 16 | Yes | Health behaviours and diet |
| TotalAlcUnits16 | TOTAL UNITS OF ALCOHOL IN PAST WEEK | 16 | Yes | Health behaviours and diet |
| VegReligion16 | VEGETARIAN BECAUSE OF RELIGIOUS REASONS | 16 | Yes | Religion, spirtuality and wider culture |
| ParentHitYou16 | WHICH PARENT HIT YOU? | 16 | Yes | ACE |
| RelBornInto16 | WHAT RELIGION WERE YOU BORN INTO? | 16 | Yes | Religion, spirtuality and wider culture |
| RelImportant16 | IS RELIGION IMPORTANT PART OF YOUR LIFE? | 16 | Yes | Religion, spirtuality and wider culture |
| RelLuckyBeleifs16 | RELIGIOUS PEOPLE LUCKY TO HAVE BELIEFS | 16 | Yes | Religion, spirtuality and wider culture |
| RelOldFashioned16 | RELIGIOUS PEOPLE ARE OLD FASHIONED | 16 | Yes | Religion, spirtuality and wider culture |
| RelValuedSoc16 | RELIGIOUS PEOPLE VALUED MEMBERS OF SOC. | 16 | Yes | Religion, spirtuality and wider culture |
| RelMisguided16 | RELIGIOUS PEOPLE ARE MISGUIDED | 16 | Yes | Religion, spirtuality and wider culture |
| RelHelpTrouble16 | RELIGIOUS PEOPLE HELP YOU IF IN TROUBLE | 16 | Yes | Religion, spirtuality and wider culture |
| RelNoDiff16 | RELIGIOUS PEOPLE NO DIFFERENT FROM REST | 16 | Yes | Religion, spirtuality and wider culture |
| NoCig16 | NUMBER OF CIGARETTES SMOKED IN A WEEK | 16 | Yes | Health behaviours and diet |
| NoisyNeighbours16 | IN AREA NOISY NEIGHBOURS OR LOUD PARTIES | 16 | Yes | Neighbourhood, physical environments and health care systems |
| Grafiti16 | IN AREA GRAFFITI ON WALLS OR BUILDINGS | 16 | Yes | Neighbourhood, physical environments and health care systems |
| Teenagersstreet16 | IN AREA TEENAGERS HANGING ROUND STREETS | 16 | Yes | Neighbourhood, physical environments and health care systems |
| DrunksStreet16 | IN AREA DRUNKS OR TRAMPS ON STREETS | 16 | Yes | Neighbourhood, physical environments and health care systems |
| RubbishStreet16 | IN AREA LOTS OF RUBBISH LYING ABOUT | 16 | Yes | Neighbourhood, physical environments and health care systems |
| UnfairlyTreaTRel16 | UNFAIRLY TREATED BECAUSE OF RELIGION | 16 | Yes | Religion, spirtuality and wider culture |
| U10SexualApproach16 | UNDER 10YRS WHEN SEXUAL APPROACH MADE | 16 | Yes | ACE |
| SexualAppr10_15 | AGED 10-15YRS WHEN SEXUAL APPROACH MADE | 16 | Yes | ACE |
| SexualApproach1yr16 | SEXUAL APPROACH MADE IN PAST YEAR | 16 | Yes | ACE |

| | | | | |
|---|---|---|---|---|
| NoSexualApproach1yr16 | NO. TIMES PAST YR UNWELCOME SEX APPROACH | 16 | Yes | ACE |
| NoAlcoholRel16 | DON'T DRINK ALCOHOL - RELIGION FORBIDS | 16 | Yes | Religion, spirtuality and wider culture |
| HealthyDiet16 | KNOW HOW TO GET A HEALTHY DIET | 16 | Yes | Health behaviours and diet |
| SportCentreFarAway16 | SPORT-COMMUNITY CENTRE TOO FAR AWAY | 16 | Yes | Neighbourhood, physical environments and health care systems |
| ActReligion16 | OTHER ACTIV. CONNECTED TO YOUR RELIGION | 16 | Yes | Religion, spirtuality and wider culture |
| MotherIllness16 | MOTHER ILLNESS OR DISABLEMENT SINCE 10YR | 16 | Yes | Transgenerational impact of parent health and health behaviours |
| FatherIllness16 | FATHER ILLNESS OR DISABLEMENT SINCE 10YR | 16 | Yes | Transgenerational impact of parent health and health behaviours |
| HusbandSmoke16 | DOES HUSBAND SMOKE AT ALL? | 16 | Yes | Transgenerational impact of parent health and health behaviours |
| MotherSmoke16 | SELF (MOTHER) SMOKE AT ALL? | 16 | Yes | Transgenerational impact of parent health and health behaviours |
| DescNeighbourhood16 | DESCRIPTION OF NEIGHBOURHOOD | 16 | Yes | Neighbourhood, physical environments and health care systems |
| CMEatPeasGreens16 | TEENAGER EATS PEAS/GREEN BEANS | 16 | Yes | Health behaviours and diet |
| CMEatOtherGreens16 | TEENAGER EATS OTHER GREEN VEGATABLES | 16 | Yes | Health behaviours and diet |
| CMEatRootVeg16 | TEENAGER EATS ROOT VEGATABLES | 16 | Yes | Health behaviours and diet |
| CMEatGreenSalad16 | TEENAGER EATS GREEN SALAD | 16 | Yes | Health behaviours and diet |
| CMEatFruit16 | TEENAGER EATS FRESH FRUIT | 16 | Yes | Health behaviours and diet |
| DietForReligion16 | IS SPECIAL DIET FOR RELIGION/CULTURE? | 16 | Yes | Religion, spirtuality and wider culture |
| CMAlcohol16 | HOW OFTEN TEENAGER HAS ALCOHOLIC DRINK | 16 | Yes | Health behaviours and diet |
| FatherAlcohol16 | HOW OFTEN HUSBAND HAS ALCOHOLIC DRINK | 16 | Yes | Health behaviours and diet |
| MotherAlcohol16 | HOW OFTEN MOTHER HAS ALCOHOLIC DRINK | 16 | Yes | Health behaviours and diet |
| CMStayHealthy16 | DOES TEENAGER DO THINGS TO KEEP HEALTHY? | 16 | Yes | Health behaviours and diet |
| MotherStayHealthy16 | DOES MOTHER DO THINGS TO KEEP HEALTHY? | 16 | Yes | Health behaviours and diet |
| HusbandStayHealthy16 | DOES HUSBAND DO THINGS TO KEEP HEALTHY? | 16 | Yes | Health behaviours and diet |

| | | | | |
|---|---|---|---|---|
| TriedSolvents16 | EVER TRIED SNIFFING GLUE/SOLVENTS? | 16 | Yes | Health behaviours and diet |
| TriedUppers16 | HAVE YOU EVER TRIED TAKING UPPERS? | 16 | Yes | Health behaviours and diet |
| TriedDowners16 | HAVE YOU EVER TRIED TAKING DOWNERS? | 16 | Yes | Health behaviours and diet |
| TriedCannabis16 | HAVE YOU EVER TRIED TAKING CANNABIS? | 16 | Yes | Health behaviours and diet |
| TriedCocaine16 | HAVE YOU EVER TRIED TAKING COCAINE? | 16 | Yes | Health behaviours and diet |
| TriedSemeron16 | HAVE YOU EVER TRIED TAKING SEMERON? | 16 | Yes | Health behaviours and diet |
| TriedHeroin16 | HAVE YOU EVER TRIED TAKING HEROIN? | 16 | Yes | Health behaviours and diet |
| Neighbourhood16 | WHAT ARE PEOPLE LIKE IN NEIGHBOURHOOD? | 16 | Yes | Neighbourhood, physical environments and health care systems |
| WalkAloneDark16 | EVER WALK ALONE IN YOUR AREA AFTER DARK? | 16 | Yes | Neighbourhood, physical environments and health care systems |
| SafeWalkAlone16 | IF WALK ALONE HOW SAFE WOULD YOU FEEL? | 16 | Yes | Neighbourhood, physical environments and health care systems |
| DescArea16 | DESCRIBE HOUSES-FLATS IN YOUR AREA | 16 | Yes | Neighbourhood, physical environments and health care systems |
| ParentalMalaise16 | TOTAL MALAISE SCORE GROUPED ( AGE 16) | 16 | Yes | Transgenerational impact of parent health and health behaviours |
| CMMalaise16 | TOTAL MALAISE SCORE (22 QUESTIONS) GROUPED | 16 | Yes | Child health including check-ups and screening |

**Supplementary Table 2. Variables identified from the data audit in NCDS**

| **Variable** | **Description** | **Sweep of data collection** | **Supplementary variable** | **Domain** |
|---|---|---|---|---|
| ncdsid | SERIAL NUMBER | N/A | N/A | N/A |
| n0region | REGION AT PMS (1958) - BIRTH | Birth | No | Prenatal, antenatal, neonatal and birth |

| | | | | |
|---|---|---|---|---|
| MatAge | MOTHER'S AGE LAST BIRTHDAY,IN YEARS | Birth | No | Prenatal, antenatal, neonatal and birth |
| Matweight | MOTHER'S WEIGHT IN STONES,1958 | Birth | No | Prenatal, antenatal, neonatal and birth |
| AnteVisits | TOTAL NUMBER OF ANTENATAL VSITS | Birth | No | Prenatal, antenatal, neonatal and birth |
| MatSmoke | SMOKING DURING PREGNANCY | Birth | No | Prenatal, antenatal, neonatal and birth |
| Parity | PARITY | Birth | No | Prenatal, antenatal, neonatal and birth |
| LabDur | DURATION OF LABOUR-1ST STAGE:HOURS | Birth | No | Prenatal, antenatal, neonatal and birth |
| FoetDiss | FOETAL DISTRESS | Birth | No | Prenatal, antenatal, neonatal and birth |
| Birthweight | WEIGHT OF BABY IN OUNCES | Birth | No | Prenatal, antenatal, neonatal and birth |
| BirthSpace | INTERVAL BETWEEN THIS BIRTH AND LAST | Birth | No | Prenatal, antenatal, neonatal and birth |
| Breastfed | BREAST FED PARTIALLY OR WHOLLY | Birth | No | Prenatal, antenatal, neonatal and birth |
| POD | PLACE OF DELIVERY | Birth | No | Prenatal, antenatal, neonatal and birth |
| PLANC | PLACE OF ANTENATAL CARE | Birth | No | Prenatal, antenatal, neonatal and birth |
| ABNORM00 | NO OBSTETRIC, PREGNANCY ABNORMALITY | Birth | No | Prenatal, antenatal, neonatal and birth |
| AD2HOSP | ADMISSION TO HOSPITAL | Birth | No | Prenatal, antenatal, neonatal and birth |
| RESUS | RESUSCITATION | Birth | No | Prenatal, antenatal, neonatal and birth |
| ILLNESS | BABY'S ILLNESS | Birth | No | Prenatal, antenatal, neonatal and birth |
| gmeasles11a | GERMAN MEASLES DERIVED VARIABLE | 11 | No | Child health including check-ups and screening |
| mumps11a | MUMPS DERIVED VARIABLE | 11 | No | Child health including check-ups and screening |
| chickenpox11a | CHICKEN POX DERIVED VARIABLE | 11 | No | Child health including check-ups and screening |
| whoopingc11a | WHOOPING COUGH DERIVED VARIABLE | 11 | No | Child health including check-ups and screening |
| scarletfever11a | SCARLET FEVER DERIVED VARIABLE | 11 | No | Child health including check-ups and screening |
| rheumaticfev11 | RHEUMATIC FEVER DERIVED VARIABLE | 11 | No | Child health including check-ups and screening |
| hepatitis11 | HEPATITIS DERIVED VARIABLE | 11 | No | Child health including check-ups and screening |
| meningitis11 | MENINGITIS DERIVED VARIABLE | 11 | No | Child health including check-ups and screening |
| tb11 | TB DERIVED VARIABLE | 11 | No | Child health including check-ups and screening |
| infecdis11a | INFECTIOUS DISEASE DERIVED VARIABLE | 11 | No | Child health including check-ups and screening |
| asthma11b | EVER ASTHMA | 11 | No | Child health including check-ups and screening |
| bronchitis11b | EVER WHEEZY BRONCHITIS | 11 | No | Child health including check-ups and screening |
| hayfever11 | N PAST YEAR-ANY HAY FEVER, RHINITIS | 11 | No | Child health including check-ups and screening |

| | | | | |
|---|---|---|---|---|
| recvomiting11 | IN PAST YEAR-RECURRENT VOMITING | 11 | No | Child health including check-ups and screening |
| mouthulcers11 | IN PAST YEAR-RECURRENT MOUTH ULCERS | 11 | No | Child health including check-ups and screening |
| throatinfec11a | RECURR THROAT,EAR INFECTS REQ TRTMENT | 11 | No | Child health including check-ups and screening |
| nephritis11 | NEPHRITIS DERIVED VARIABLE | 11 | No | Child health including check-ups and screening |
| nephrosis11 | NEPHROSIS DERIVED VARIABLE | 11 | No | Child health including check-ups and screening |
| urinaryinfec11 | URINARY INFECTION DERIVED VARIABLE | 11 | No | Child health including check-ups and screening |
| poorbreathing11 | POOR BREATHING DERIVED VARIABLE | 11 | No | Child health including check-ups and screening |
| manycolds11 | MANY COLDS DERIVED VARIABLE | 11 | No | Child health including check-ups and screening |
| redeyes11 | 2 SORE, RED EYES DERIVED VARIABLE | 11 | No | Child health including check-ups and screening |
| coldhands11 | VERY COLD HANDS DERIVED VARIABLE | 11 | No | Child health including check-ups and screening |
| epilepsy11 | EPILEPY DERIVED VARIABLE | 11 | No | Child health including check-ups and screening |
| measles11 | MEASLES COMPOSITE DERIVED VARIABLE | 11 | No | Child health including check-ups and screening |
| headache11y | HEADACHES COMPOSITE DERIVED VARIABLE | 11 | No | Child health including check-ups and screening |
| abpain11y | ABDOMINAL PAIN COMPOSITE DERIVED VARIABLE | 11 | No | Child health including check-ups and screening |
| heartcondition11y | HEART CONDITION COMPOSITE DERIVED VARIABLE | 11 | No | Child health including check-ups and screening |
| eczema11y | ECZEMA COMPOSITE DERIVED VARIABLE | 11 | No | Child health including check-ups and screening |
| hernia11y | HERNIA COMPOSITE DERIVED VARIABLE | 11 | No | Child health including check-ups and screening |
| tonsilitis11y | TONSILITIS COMPOSITE DERIVED VARIABLE | 11 | No | Child health including check-ups and screening |
| somaticsymptoms11y | SCALE TOTAL SOMATIC SYMPTOMS 2P | 11 | No | Child health including check-ups and screening |
| gastrosymptoms11y | SCALE TOTAL GASTROINTESTINAL SYMPTOMS 2P | 11 | No | Child health including check-ups and screening |
| gastroillness11y | SCALE TOTAL GASTROINTESTINAL ILLNESS 2P | 11 | No | Child health including check-ups and screening |
| longstandingIll11y | SCALE TOTAL LONGSTANDING ILLNESS 2P | 11 | No | Child health including check-ups and screening |
| atopy11y | SCALE TOTAL ATOPY 2P | 11 | No | Child health including check-ups and screening |
| infecillness11 | SCALE TOTAL INFECTIOUS ILLNESS 2P | 11 | No | Child health including check-ups and screening |
| Hearing11 | CHILD ALWAYS GOOD HEARING BOTH EARS | 11 | No | Child health including check-ups and screening |
| HospAd11 | NO.OF TIMES CHLD ADMITTED TO HOSPITAL | 11 | No | Child health including check-ups and screening |
| ChildOutPatient | HAS CHILD BEEN AN OUTPATIENT | 11 | No | Child health including check-ups and screening |

| | | | | |
|---|---|---|---|---|
| PsychTreatment | PSYCHIATRIC,PSYCHOLOGICAL TREATMENT | 11 | No | Child health including check-ups and screening |
| BSAGAnxiety | TOT SCORE-BSAG ANXIETY ACCEPTNCE,CHILDN | 11 | No | Child health including check-ups and screening |
| BSAGDepression | TOTAL SCORE-BSAG DEPRESSION SYNDROME | 11 | No | Child health including check-ups and screening |
| ChildIrritable | IS CHILD IRRITABLE,QUICK TEMPERED | 11 | No | Developmental attributes |
| ChildClumsy | ANY ABNORMALITY,CLUMSINESS-MC 1:4 | 11 | No | Developmental attributes |
| WalkLine | WALKING BACKWARDS ALONG STRAIGHT LINE | 11 | No | Developmental attributes |
| StandRight | STANDING ON RIGHT FOOT 15 FOR SECONDS | 11 | No | Developmental attributes |
| StandLeft | STANDING ON LEFT FOOT FOR 15 SECONDS | 11 | No | Developmental attributes |
| HeelToe | STANDING HEEL TO TOE FOR 15 SECONDS | 11 | No | Developmental attributes |
| ChildHandControl | CHILD HAS POOR HAND CONTROL | 11 | No | Developmental attributes |
| ChildCoord | CHILD HAS POOR PHYSICAL CO-ORDINATION | 11 | No | Developmental attributes |
| InconsBeh | TOT SCORE BSAG INCONSEQUENTIAL BEHAVIOR | 11 | No | Developmental attributes |
| NervSymptoms | TOT SCORE BSAG MISC NERVOUS SYMPTOMS | 11 | No | Developmental attributes |
| GeneralKnowTR | CHILD'S GEN KNOWLDGE-TEACHER'S RATING | 11 | No | Education and health literacy |
| NumberTR | CHILD'S NUMBER WORK-TEACHER'S RATING | 11 | No | Education and health literacy |
| BookTR | CHILD'S USE BOOKS-TEACHER'S RATING | 11 | No | Education and health literacy |
| OralAbTR | CHILD'S ORAL ABILITY-TEACHER'S RATING | 11 | No | Education and health literacy |
| GraspEng | IMPERFECT GRASP OF ENGLISH | 11 | No | Education and health literacy |
| GeneralAbT | TOTAL SCORE ON GENERAL ABILITY TEST | 11 | No | Education and health literacy |
| ReadingCompT | READING COMPREHENSION TEST SCORE | 11 | No | Education and health literacy |
| MathT | MATHEMATICS TEST SCORE | 11 | No | Education and health literacy |
| CopyT | COPYING DESIGNS TEST SCORE | 11 | No | Education and health literacy |
| Sex | 0-3D SEX OF CHILD | 11 | No | Demographic |
| Region | REGION AT NCDS2 (1969) - 11 YEARS | 11 | No | Demographic |
| HHNumber | NUMBER LIVING IN CHILD'S HOUSEHOLD | 11 | No | Demographic |
| MotherEthnicity | AREA OF WORLD IN WHICH MOTHER BORN | 11 | No | Demographic |
| FatherEthnicity | AREA OF WORLD IN WHICH FATHER BORN | 11 | No | Demographic |
| LangHome | IS ENGLISH USUALLY SPOKEN AT HOME | 11 | No | Demographic |
| FamilyMoves | NO. OF FAMILY MOVES SINCE CHLDS BIRTH | 11 | No | Demographic |

| | | | | |
|---|---|---|---|---|
| ParentalSep | PARENTS SEPARATED | 11 | No | Demographic/ACE |
| ParentalDeath | PARENTAL DEATH | 11 | No | Demographic/ACE |
| MotherChronicCon | WHAT IS MUMS CHRONIC CONDITION M:C1-2 | 11 | No | Transgenerational impact of parent health and health behaviours |
| FatherChronicCon | WHAT IS DADS CHRONIC CONDITION M:C1-2 | 11 | No | Transgenerational impact of parent health and health behaviours |
| FatherWeight | FATHER'S WEIGHT IN STONES | 11 | No | Transgenerational impact of parent health and health behaviours |
| FatherHeight | FATHER'S HEIGHT IN INCHES | 11 | No | Transgenerational impact of parent health and health behaviours |
| MotherWeight | MOTHER'S WEIGHT IN STONES | 11 | No | Transgenerational impact of parent health and health behaviours |
| FatherWeight | MOTHERS HEIGHT IN INCHES | 11 | No | Transgenerational impact of parent health and health behaviours |
| HHnum | NUMBER LIVING IN CHILD'S HOUSEHOLD | 11 | No | Socioeconomics/Demographic |
| HousingTenure | TENURE OF ACCOMODATION | 11 | No | Socioeconomics/Demographic |
| NoShareBedroom | HOW MANY PEOPLE SHARE CHILDS BEDROOM | 11 | No | Socioeconomics/Demographic |
| HHAmenities | ACCESS TO HOUSEHOLD AMENITIES | 11 | No | Socioeconomics/Demographic |
| NumberPerRoom | NUMBER OF PERSONS PER ROOM | 11 | No | Socioeconomics/Demographic |
| FatherSC | SOCIAL CLASS OF FATHER OR MALE HEAD (GRO 1966) | 11 | No | Socioeconomics/Demographic |
| FatherUnemploy | FATHER,MALE HEAD'S OCCUPATION | 11 | No | Socioeconomics/Demographic |
| MotherRecSEG | MOTHERS'S MOST RECENT WORK AND SEG (GRO 1966) | 11 | No | Socioeconomics/Demographic |
| FreeSchoolMeals | DOES ANY CHILD GET FREE SCHOOL MEALS | 11 | No | Socioeconomics/Demographic |
| FinancialHardship | SERIOUS FINANCIAL HARDSHIP LAST YR | 11 | No | Socioeconomics/Demographic |
| MotherFig | CHILD'S MOTHER FIGURE | 11 | No | Demographic/ACE |
| FatherFig | CHILD'S FATHER FIGURE | 11 | No | Demographic/ACE |
| LACare | HAS CHILD EVER BEEN IN LA CARE | 11 | No | ACE |
| VolServCare | HAS CHILD BEEN IN VOL SERVICE CARE | 11 | No | ACE |
| ChildDisob | IS CHILD DISOBEDIENT AT HOME | 11 | No | Developmental attributes/Parental family factors and parental ability to care for child |
| ParSchLeaveAge | PARENTAL HOPES CHILD'S SCHOOL LEAVING | 11 | No | Parental family factors and parental ability to care for child |
| ParFurtherEd | PARS WANT FURTHR EDUC,TRAIN FOR CHLD | 11 | No | Parental family factors and parental ability to care for child |

| | | | | |
|---|---|---|---|---|
| MotherWalk | DOES MUM TAKE CHILD FOR WALKS,VISITS | 11 | No | Parental family factors and parental ability to care for child |
| FatherWalk | DOES DAD TAKE CHILD FOR WALKS,VISITS | 11 | No | Parental family factors and parental ability to care for child |
| FatherMangChild | DADS ROLE IN MANAGEMENT OF CHILD | 11 | No | Parental family factors and parental ability to care for child |
| FatherIntrEd | FATHERS' INTEREST IN CHILDS EDUCATION | 11 | No | Parental family factors and parental ability to care for child |
| MotherIntrEd | MOTHERS' INTEREST IN CHILDS EDUCATION | 11 | No | Parental family factors and parental ability to care for child |
| PlayAreas | IS MUM HAPPY WITH PLAY AREAS NEARBY | 11 | No | Neighbourhood, physical environments and health care systems |
| PublicParks | USE OF PUBLIC PARKS, ETC IN LAST 12M | 11 | No | Neighbourhood, physical environments and health care systems |
| RecreationGround | USE OF RECREATION GRND, ETC LAST 12M | 11 | No | Neighbourhood, physical environments and health care systems |
| Overeating7 | OVEREATING-REPORTED BY MOTHER | 7 | Yes | Health behaviours and diet |
| ContactProbation7 | FAMILY CONTACT-PROBATION OFFICER | 7 | Yes | ACE |
| FamDiffMentalll7 | FAM DIFFICLTS-MENTAL ILLNESS,NEUROSIS | 7 | Yes | ACE/Transgenerational impact of parent health and health behaviours |
| FamDiffMentaSub7 | FAM DIFFICULTIES-MENTAL SUBNORMALITY | 7 | Yes | ACE/Transgenerational impact of parent health and health behaviours |
| FamDiffDomesticTen7 | FAMILY DIFFICULTIES-DOMESTIC TENSION | 7 | Yes | ACE |
| FamDiffAlcohol7 | FAMILY DIFFICULTIES-ALCOHOLISM | 7 | Yes | ACE/Transgenerational impact of parent health and health behaviours |
| MotherSmoking16 | NO. OF CIGARETTES MUM SMOKES PER DAY | 16 | Yes | Transgenerational impact of parent health and health behaviours |
| FatherSmoking16 | NO. OF CIGARETTES DAD SMOKES PER DAY | 16 | Yes | Transgenerational impact of parent health and health behaviours |
| AnyEatingDiff7 | IS THERE ANY EATING DIFFICULTY | 7 | Yes | Health behaviours and diet |
| TypeEatDisorder7 | TYPE OF EATING DIFFICULTY M:C 1-2 | 7 | Yes | Health behaviours and diet |
| MotherIllness16 | MOTHER-DIAGNOSIS OF ILLNESS | 16 | Yes | Transgenerational impact of parent health and health behaviours |
| FatherIllness16 | FATHER-DIAGNOSIS OF ILLNESS | 16 | Yes | Transgenerational impact of parent health and health behaviours |
| AptitudeSport16 | APTITUDE FOR SPORTS & GAMES-STDY CHLD | 16 | Yes | Health behaviours and diet |

| | | | | |
|---|---|---|---|---|
| SatisfyPlaceMeet16 | SATISFACTION - PLACES TO MEET IN AREA | 16 | Yes | Neighbourhood, physical environments and health care systems |
| SatisfySportFac16 | SATISFIED-SPORTING FACILITIES IN AREA | 16 | Yes | Neighbourhood, physical environments and health care systems |
| NoCigsAWeek16 | NO. OF CIGARETTES SMOKED PER WEEK | 16 | Yes | Health behaviours and diet |
| HowLongSinceAlcohol16 | HOW LONG SINCE CHILD DRANK ALCOHOL | 16 | Yes | Health behaviours and diet |
| AlcoholLastWeek16 | NO. AND TYPE DRINKS LAST WEEK-MC 1:3 | 16 | Yes | Health behaviours and diet |
| Childactive7 | CHILD NORMALLY ACTIVE-MUMS VIEW | 16 | Yes | Health behaviours and diet |
| Sport11 | PUPIL TAKES PART IN SPORT OUT OF SCHOOL | 11 | No | Health behaviours and diet |
| Outdoorsport16 | HOW OFTEN PLAYS OUTDOOR GAMES & SPORT | 16 | Yes | Health behaviours and diet |
| Indoorsport16 | HOW OFTEN PLAYS INDOOR GAMES & SPORT | 16 | Yes | Health behaviours and diet |

**Supplementary Table 3. Variables identified from the data audit in ACONF**

| **Variable** | **Description** | **Sweep of data collection** | **Supplementary variable** | **Domain** |
|---|---|---|---|---|
| Rs001 | Sex | Reading survey | N/A | Demographic |
| M_matage | Maternal age | Reading survey | N/A | Demographic |
| Rs004 | Place of birth | Reading survey | N/A | Demographic |
| Rs007 | Population size | Reading survey | N/A | Demographic |
| M_numsib | Number of siblings | Reading survey | N/A | Demographic |
| Rs035 | Family size | Reading survey | N/A | Demographic |
| Rs041 | Marital status at birth | Reading survey | N/A | Demographic |
| Fs101 | Any step children | Family survey | N/A | Demographic |
| Rs008 | Mobility | Reading survey | N/A | Socioeconomic |
| Rs009 | Class grade | Reading survey | N/A | Socioeconomic |
| Rs014 | Number of schools attended | Reading survey | N/A | Socioeconomic |
| Rs027 | Area of residence | Reading survey | N/A | Socioeconomic |
| Rs032 | Father social class | Reading survey | N/A | Socioeconomic |
| Rs041 | Mother occupation | Reading survey | N/A | Socioeconomic |
| Rs042 | Wife education | Reading survey | N/A | Socioeconomic |

| | | | | |
|---|---|---|---|---|
| Fs106 | Mother school leaving age | Family survey | N/A | Socioeconomic |
| Sch_mate | Mother further education | Family survey | N/A | Socioeconomic |
| Fs202 | Housing tenure | Family survey | N/A | Socioeconomic |
| Fs203 | Number of rooms | Family survey | N/A | Socioeconomic |
| Fs212 | Household size | Family survey | N/A | Socioeconomic |
| Fs374 | Father unemployment | Family survey | N/A | Socioeconomic |
| C_phhcon | Percentage renting from council | Reading survey | N/A | Neighbourhood, physical environments and health care systems |
| C_phhown | Percentage of owner-occupied houses | Reading survey | N/A | Neighbourhood, physical environments and health care systems |
| C_phhhtn | Percentage of houses with hot water | Reading survey | N/A | Neighbourhood, physical environments and health care systems |
| C_phcldn | Percentage of houses with cold water | Reading survey | N/A | Neighbourhood, physical environments and health care systems |
| C_phbthn | Percentage of households with no fixed bath | Reading survey | N/A | Neighbourhood, physical environments and health care systems |
| C_phhwc | Percentage of households with shared WC | Reading survey | N/A | Neighbourhood, physical environments and health care systems |
| Fs054 | Behavioural problems | Family survey | N/A | Developmental attributes |
| Fs135 | Rutter scale A | Family survey | N/A | Developmental attributes |
| Fs219 | Sociability of child | Family survey | N/A | Developmental attributes |
| Rs173 | Anti-social score scale B | Reading survey | N/A | Developmental attributes |
| Rs174 | Neurotic score scale B | Reading survey | N/A | Developmental attributes |
| Rs175 | Total score scale B | Reading survey | N/A | Developmental attributes |
| Rs176 | Neurotic/anti-social rating | Reading survey | N/A | Developmental attributes |

| | | | | |
|---|---|---|---|---|
| Rs030 | Absences from school for health reasons | Reading survey | N/A | Child health including check-ups and screening |
| Rs109 | Number of medical exams | Reading survey | N/A | Child health including check-ups and screening |
| Rs113-rs115 | Height | Reading survey | N/A | Child health including check-ups and screening |
| Rs117-rs118 | Weight | Reading survey | N/A | Child health including check-ups and screening |
| Fs015 | Hospital admissions | Family survey | N/A | Child health including check-ups and screening |
| Fs025 | Asthma | Family survey | N/A | Child health including check-ups and screening |
| Fs026 | Measles | Family survey | N/A | Child health including check-ups and screening |
| Fs027 | Hay fever | Family survey | N/A | Child health including check-ups and screening |
| Fs028 | Whooping cough | Family survey | N/A | Child health including check-ups and screening |
| Fs029 | Eczema | Family survey | N/A | Child health including check-ups and screening |
| Fs030 | Convulsions/fits | Family survey | N/A | Child health including check-ups and screening |
| Fs031 | Meningitis | Family survey | N/A | Child health including check-ups and screening |
| Fs032 | Cold | Family survey | N/A | Child health including check-ups and screening |
| Fs033 | Sore throat | Family survey | N/A | Child health including check-ups and screening |
| Fs034 | Cough | Family survey | N/A | Child health including check-ups and screening |
| Fs035 | Bronchitis | Family survey | N/A | Child health including check-ups and screening |
| Fs036 | Hives | Family survey | N/A | Child health including check-ups and screening |
| Fs037 | Stomach ache | Family survey | N/A | Child health including check-ups and screening |

| | | | | |
|---|---|---|---|---|
| Fs038 | Ear ache | Family survey | N/A | Child health including check-ups and screening |
| Fs039 | Other frequent illness | Family survey | N/A | Child health including check-ups and screening |
| Fs040 | Shortness of breath | Family survey | N/A | Child health including check-ups and screening |
| Fs041 | Wheezy chest | Family survey | N/A | Child health including check-ups and screening |
| Fs051 | Physical health | Family survey | N/A | Child health including check-ups and screening |
| Fs052 | Mental and emotional health | Family survey | N/A | Child health including check-ups and screening |
| Fs073 | Nursery school attendance | Family survey | N/A | Education and health literacy |
| Rs180 | Intelligence test at age 7 | Reading survey | N/A | Education and health literacy |
| Rs181 | Intelligence test at age 9 | Reading survey | N/A | Education and health literacy |
| Rs094 | Type of school | Reading survey | N/A | Education and health literacy |
| Rs179 | Degree of over/under achievement at age 11 | Reading survey | N/A | Education and health literacy |
| Sch_iq7 | School IQ at age 7 | Family survey | N/A | Education and health literacy |
| Sch_iq9 | School IQ at age 9 | Reading survey | N/A | Education and health literacy |
| Rs074 | Mother physical grade | Reading survey | N/A | Prenatal, antenatal, neonatal and birth |
| Rs090 | Child physical grade | Reading survey | N/A | Prenatal, antenatal, neonatal and birth |
| Rs076 | Haemorrhage complications | Reading survey | N/A | Prenatal, antenatal, neonatal and birth |
| Rs080 | Type of delivery | Reading survey | N/A | Prenatal, antenatal, neonatal and birth |
| Rs081 | Length of labour | Reading survey | N/A | Prenatal, antenatal, neonatal and birth |

| | | | | |
|---|---|---|---|---|
| Rs082 | Caesarean | Reading survey | N/A | Prenatal, antenatal, neonatal and birth |
| Rs083-rs084 | Week of antenatal visit | Reading survey | N/A | Prenatal, antenatal, neonatal and birth |
| Rs085-rs086 | Gestational length | Reading survey | N/A | Prenatal, antenatal, neonatal and birth |
| Rs087 | Birth weight | Reading survey | N/A | Prenatal, antenatal, neonatal and birth |
| Rs075 | Pre eclamptic toxaemia complications | Reading survey | N/A | Prenatal, antenatal, neonatal and birth |

**Supplementary Figure 1. PCA analysis mapping mutually exclusive variables within each domain based on similar characteristics in ACONF.**

*PDF Document*

**Supplementary Figure 2. Using PCA analysis to map mutually exclusive variables within each domain based on similar characteristics in NCDS.**

*PDF Document*

**Supplementary Figure 3. Using PCA analysis to map mutually exclusive variables within each domain based on similar characteristics in NCDS.**

*PDF Document*
