## Supplementary figures and images for "Mapping domains of early-life determinants of future multimorbidity across three UK longitudinal cohort studies"

### Supplementary Fig 1

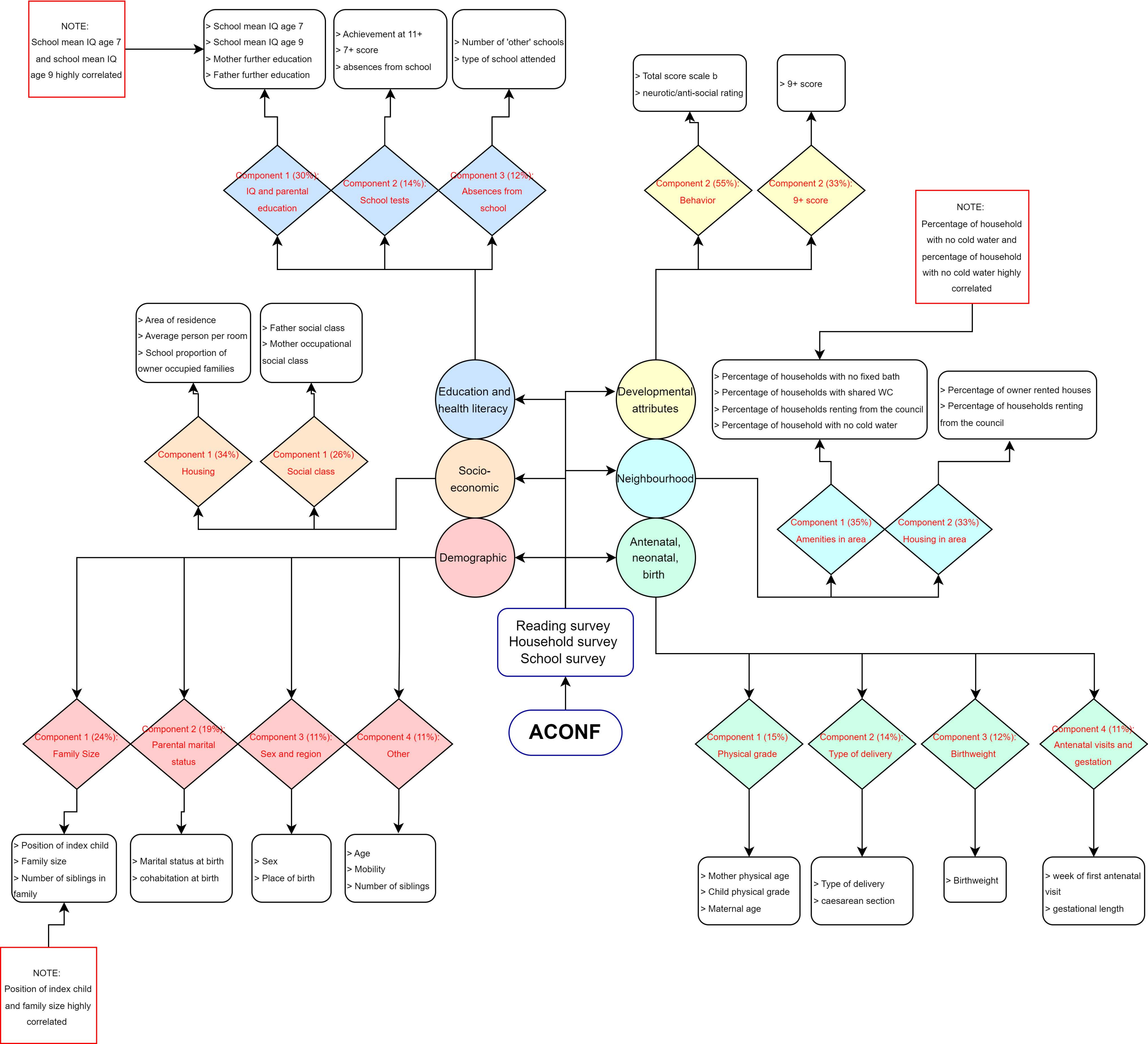

### Supplementary Fig 2

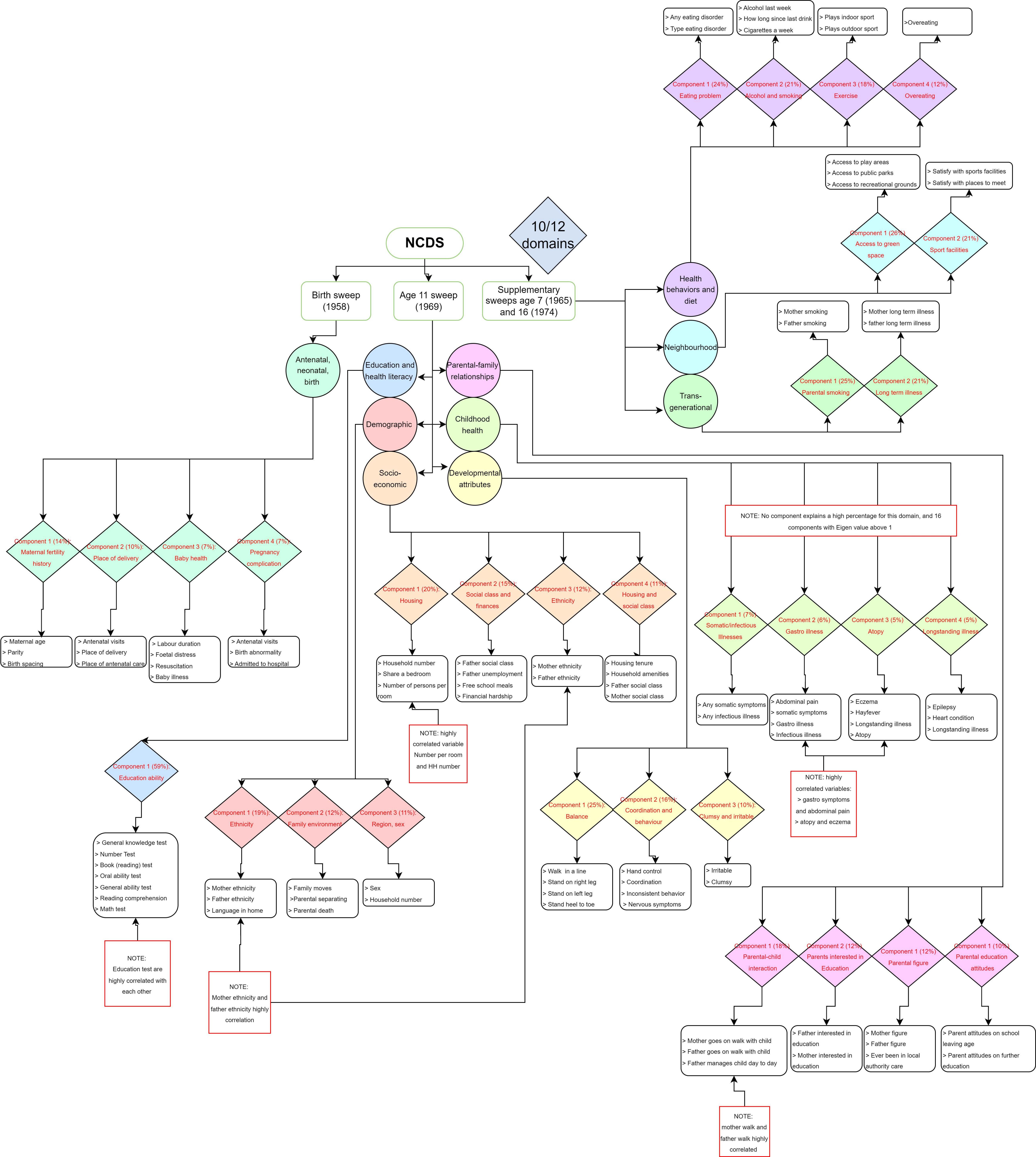

### Supplementary Fig 3

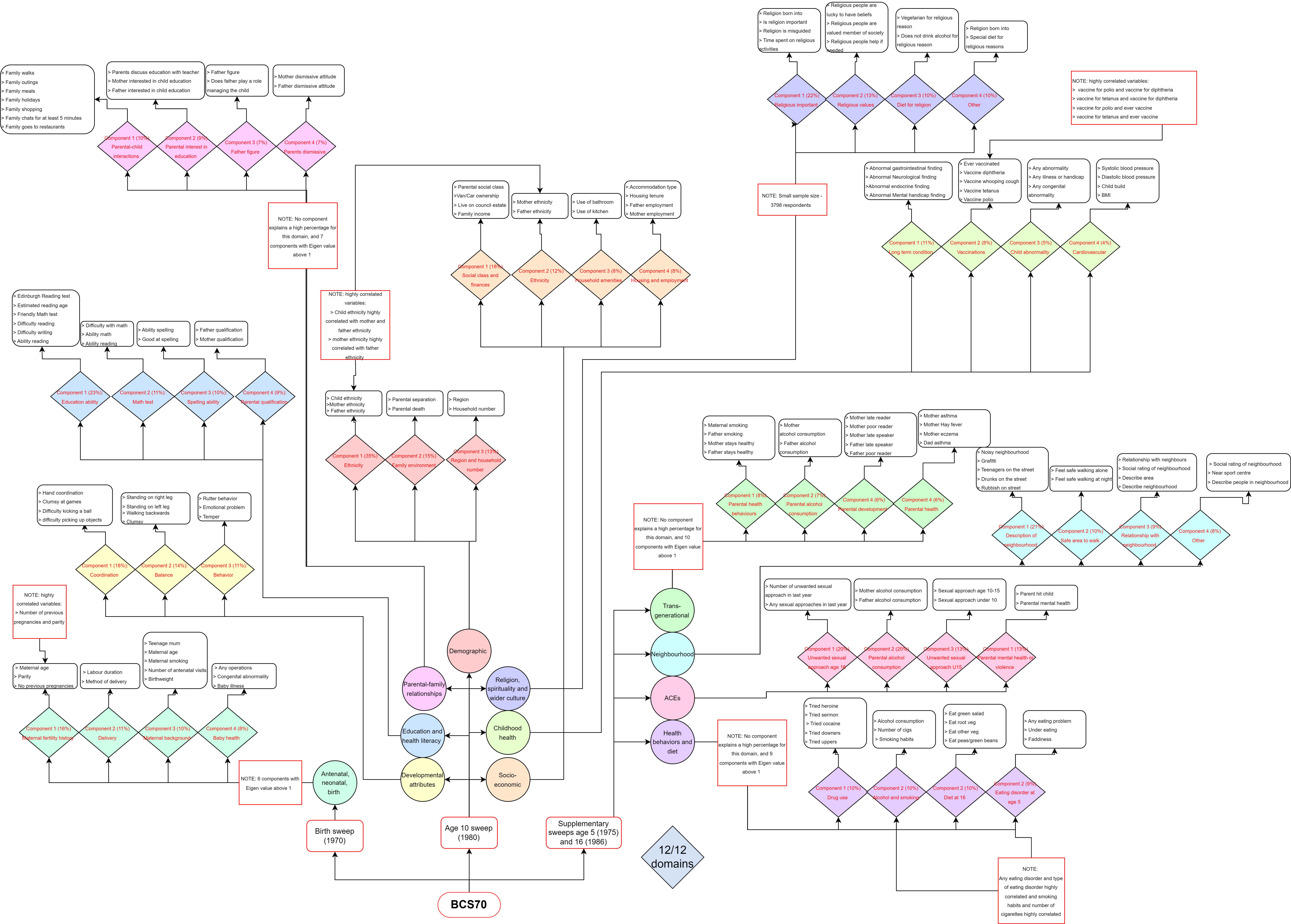
